# New-Onset IgG Autoantibodies in Hospitalized Patients with COVID-19

**DOI:** 10.1101/2021.01.27.21250559

**Authors:** Sarah Esther Chang, Allan Feng, Wenzhao Meng, Sokratis A. Apostolidis, Elisabeth Mack, Maja Artandi, Linda Barman, Kate Bennett, Saborni Chakraborty, Iris Chang, Peggie Cheung, Sharon Chinthrajah, Shaurya Dhingra, Evan Do, Amanda Finck, Andrew Gaano, Reinhard Geßner, Heather M. Giannini, Joyce Gonzalez, Sarah Greib, Margrit Gündisch, Alex Ren Hsu, Alex Kuo, Monali Manohar, Rong Mao, Indira Neeli, Andreas Neubauer, Oluwatosin Oniyide, Abigail Elizabeth Powell, Rajan Puri, Harald Renz, Jeffrey M. Schapiro, Payton A Weidenbacher, Rich Wittman, Neera Ahuja, Ho-Ryun Chung, Pras Jagannathan, Judith James, Peter S. Kim, Nuala J. Meyer, Kari Nadeau, Marko Radic, William H. Robinson, Upinder Singh, Taia T. Wang, E. John Wherry, Chrysanthi Skevaki, Eline T. Luning Prak, PJ Utz

## Abstract

Coronavirus Disease 2019 (COVID-19), caused by Severe Acute Respiratory Syndrome Coronavirus-2 (SARS-CoV-2), is associated with a wide range of clinical manifestations, including autoimmune features and autoantibody production. We developed three different protein arrays to measure hallmark IgG autoantibodies associated with Connective Tissue Diseases (CTDs), Anti-Cytokine Antibodies (ACA), and anti-viral antibody responses in 147 hospitalized COVID-19 patients in three different centers. Autoantibodies were identified in approximately 50% of patients, but in <15% of healthy controls. When present, autoantibodies largely targeted autoantigens associated with rare disorders such as myositis, systemic sclerosis and CTD overlap syndromes. Anti-nuclear antibodies (ANA) were observed in ∼25% of patients. Patients with autoantibodies tended to demonstrate one or a few specificities whereas ACA were even more prevalent, and patients often had antibodies to multiple cytokines. Rare patients were identified with IgG antibodies against angiotensin converting enzyme-2 (ACE-2). A subset of autoantibodies and ACA developed *de novo* following SARS-CoV-2 infection while others were transient. Autoantibodies tracked with longitudinal development of IgG antibodies that recognized SARS-CoV-2 structural proteins such as S1, S2, M, N and a subset of non-structural proteins, but not proteins from influenza, seasonal coronaviruses or other pathogenic viruses. COVID-19 patients with one or more autoantibodies tended to have higher levels of antibodies against SARS-CoV-2 Nonstructural Protein 1 (NSP1) and Methyltransferase (ME). We conclude that SARS-CoV-2 causes development of new-onset IgG autoantibodies in a significant proportion of hospitalized COVID-19 patients and are positively correlated with immune responses to SARS-CoV-2 proteins.

## Introduction

Coronavirus Disease 2019 (COVID-19), caused by Severe Acute Respiratory Syndrome Coronavirus-2 (SARS-CoV-2) infection, is associated with many different clinical features that are commonly found in autoimmune diseases, including arthralgias, myalgias, fatigue, sicca, and rashes^1-3^. Less common manifestations of autoimmunity have also been observed in COVID-19 patients, including thrombosis, myositis, myocarditis, arthritis, encephalitis, and vasculitis^3^. These clinical observations, and the increasing proportion of “recovered” patients with persistent post-COVID-19 symptoms (so-called “long haulers”, or “long COVID”) suggest that inflammation in response to SARS-CoV-2 infection promotes tissue damage in the acute phase and potentially some of the long-term sequelae^4-6^.

Autoantibodies, a hallmark of most but not all autoimmune disorders, have been described in COVID-19 patients. In the earliest report, approximately half of hospitalized patients at an academic hospital in Greece had high levels of serum autoantibodies, often associated with clinical findings such as rashes, thrombosis, and vasculitis^7^. Serum anti-nuclear antibodies (ANA) were detectable in approximately one third of patients^7^. Woodruff *et al.* reported that 23 of 48 (44%) of critically-ill COVID-19 patients have positive ANA tests^8,9^. Zuo described an even higher prevalence of thrombogenic autoantibodies, reporting that up to 52% of hospitalized COVID-19 patients have anti- phospholipid antibodies. They further showed that autoantibodies have the capacity to cause clots in mouse models^10^. In a large autoantibody screen, Gruber *et al.* demonstrated that Multisystem Inflammatory Syndrome in Children (MIS-C) patients develop autoantibodies, including autoantibodies against the lupus antigen SSB/La^11^. SSB/Ro autoantibodies have also been described^12^. The apparent link between clinical manifestations resembling those seen in patients with classifiable autoimmune diseases, and those observed in COVID-19 patients, has prompted searches for candidate target autoantigens that may be useful for diagnosis and for improving understanding of COVID-19 pathogenesis. The molecular targets of autoantibodies in individual patients with COVID-19 are largely unknown, as are their associations with anti-viral immune responses, and the timing of their appearance in regard to infection with SARS-CoV-2.

We hypothesized that SARS-CoV-2 induces the production of antibodies against autoantigens and cytokines/chemokines *de novo*, and these correlate with anti-viral responses. We assembled three different custom bead-based protein arrays to measure IgG antibodies found in CTDs, ACA, and anti-viral responses in 197 COVID-19 samples. Samples were obtained from 147 hospitalized patients infected by SARS-CoV-2, some of which were collected longitudinally, in three geographically distinct locations. Our results demonstrate that a large cadre of autoantigens are targeted by circulating antibodies in a substantial proportion of hospitalized patients with COVID-19, but less commonly in uninfected healthy controls (HC). Our studies confirm emerging reports of IgG autoantibodies in hospitalized COVID-19 patients and demonstrate that a significant subset of patients develop new-onset autoantibodies that could place them at risk for progression to symptomatic, classifiable autoimmunity in the future.

## Results

### Anti-nuclear antibodies (ANA) are produced by one in four hospitalized COVID-19 patients

To determine if hospitalized patients with COVID-19 produce autoantibodies against prototypical autoantigens associated with systemic autoimmunity, we measured ANA using an indirect immunofluorescence assay in one of our cohorts (University of Pennsylvania). We found that seven out of 73 patients (10%) were positive at a dilution of 1:160 using a clinical-grade assay and that another 13 were weakly positive (***Supplementary Fig. 1a***). A variety of ANA patterns were observed including diffuse, speckled and nucleolar (***Supplementary Fig. 1b and 1c***). One patient exhibited cytoplasmic staining but was negative for nuclear staining. Given the finding of positive and weakly positive ANAs, we measured dsDNA antibodies. Only one individual out of 73 tested was positive for dsDNA antibodies at a dilution of 1:270, and this individual also was ANA positive with a speckled pattern (***Supplementary Fig. 2***). Since several patients who were severely or critically ill had thromboembolic and vascular events, we also analyzed the same 73 patients for Myeloperoxidase (MPO) and Proteinase 3 (PR3) antibodies, as these antibodies are associated with autoimmune vasculitis. Only one individual tested positive for PR3 antibodies (***Supplementary Fig. 2***). The levels of positivity in these clinical-grade assays are in line with those of one of the authors (J.J.) who reported that 17 of 113 (15.8%) patients with positive SARS-CoV-2 serology had serum autoantibodies and/or antiphospholipid antibodies^13^. These findings prompted us to “cast a wider net” for autoantibodies using additional patients and assays that detected larger numbers of not only common, but also unusual autoantigens.

### Protein microarrays identify autoantibody targets in hospitalized COVID-19 patients

To systematically and simultaneously measure a large number of different autoantibodies in serum or plasma derived from patients acutely infected with SARS-CoV-2, we constructed a 53-plex COVID-19 Autoantigen Array (***Fig. 1, left half of panel***). The array comprised well-characterized autoantigens (***Supplementary Table 1***) across multiple rheumatologic diseases (***Supplementary Fig. 3)***. Included were prominent antigens targeted in systemic sclerosis (scleroderma, SSc, left panel; myositis and overlap syndromes, second panel); systemic lupus erythematosus (SLE), Sjögren’s syndrome, and mixed connective tissue disease (MCTD, third panel); gastrointestinal and endocrine autoimmune disorders (fourth panel); chromatin-associated antigens (fifth panel); and miscellaneous antigens, including proteins targeted in vasculitis or in which autoantibodies are postulated to be directly pathogenic (sixth panel). Most antigens have been validated in previous publications and were also validated using commercially available autoimmune disease prototype sera (***Fig. 1, bottom panel***), or using previously characterized serum from Stanford’s biobank and the Oklahoma Immune Cohort in the Oklahoma Medical Research Foundation Arthritis & Clinical Immunology Biorepository (SLE, SSc, MCTD, primary biliary cirrhosis, and other disorders, data not shown).

**Fig. 1:**
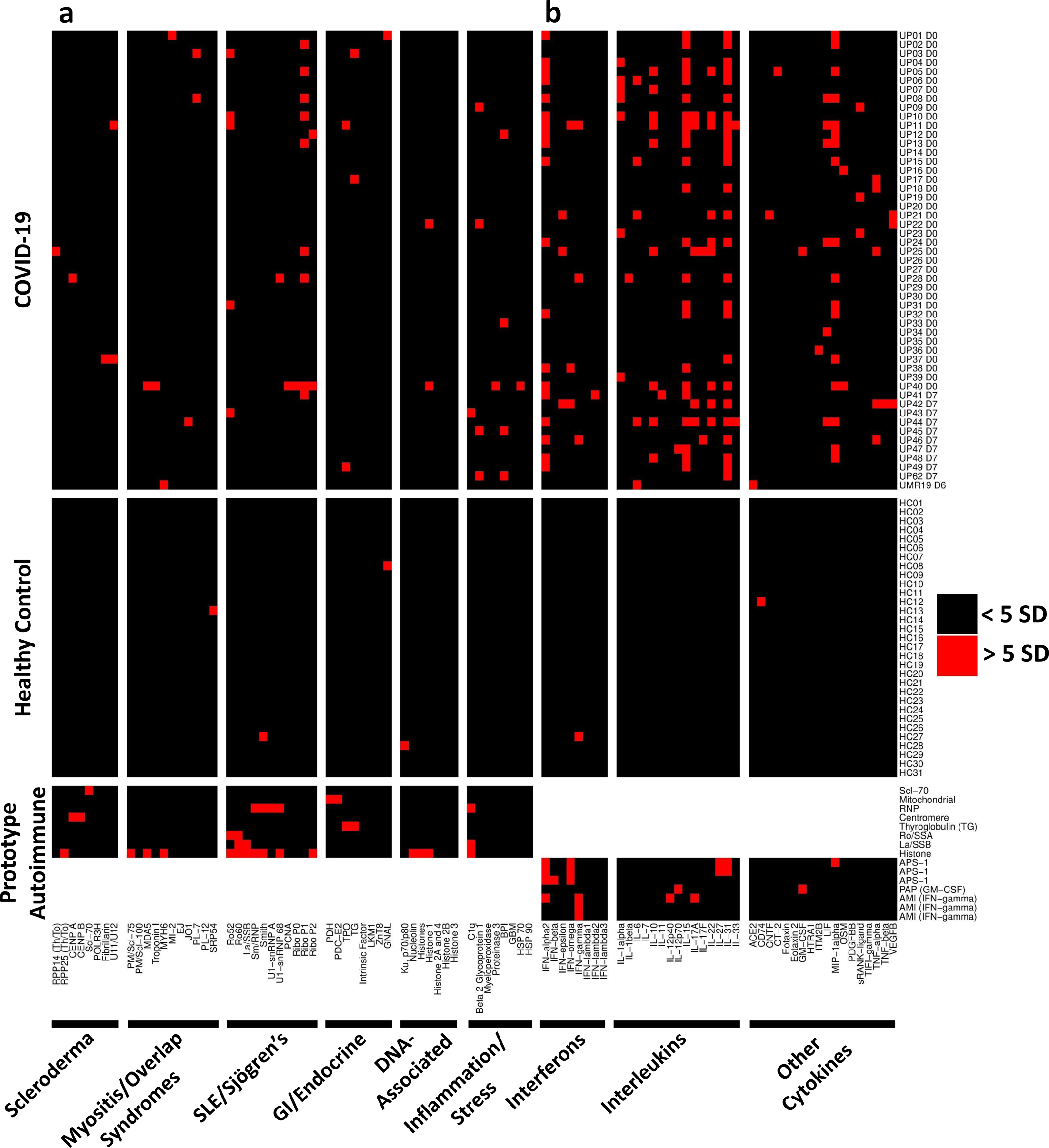
High prevalence of autoantibodies in hospitalized COVID-19 patients. **a.** Heatmap depicting serum IgG antibodies discovered using a 53-plex bead-based protein array containing the indicated autoantigens (x-axis). Autoantigens are grouped based on disease (scleroderma, myositis and overlap syndromes such as mixed connective tissue disease (MCTD), SLE/Sjögren’s, gastrointestinal and endocrine disorders), DNA-associated antigens, and antigens associated with tissue inflammation or stress responses. COVID-19 patients (top panel), HC (n=31, middle panel), and 8 prototype autoimmune disorders (bottom panel) are shown. Colors indicate autoantibodies whose MFI measurements are >5 SD (red) or <5 SD (black) above the average MFI for HC. MFIs <3,000 were excluded. **b.** Heatmap using a 41-plex array of cytokines, chemokines, growth factors, and receptors. The same samples in Panel A were also analyzed for anti-cytokine antibodies (ACA). Cytokines are grouped on the x-axis by category (interferons, interleukins, and other cytokines/growth factors/receptors). Prototype samples from patients with immunodeficiency disorders include three patients with Autoimmune Polyendocrine Syndrome Type 1 (APS1), one patient with Pulmonary Alveolar Proteinosis (PAP), and three patients with Atypical Mycobacterial Infections (AMI). Colors indicate autoantibodies whose MFI measurements are >5 SD (red) or < 5 SD (black) above the average MFI for HC. MFIs <3,000 were excluded.

We characterized 51 cross-sectional COVID-19 serum or plasma samples from patients who provided samples within seven days of hospitalization (***Fig. 1***). As expected, prototype reference samples from patients with classifiable autoimmune diseases were strongly positive for autoantibodies, recognizing 25 of the 53 arrayed proteins (***Fig. 1***, ***bottom left panel***, and ***Supplementary Fig. 3***). Serum from only four HC recognized a single autoantigen each (signal recognition particle 54, SRP 54; Smith/ribonuclear protein, Sm/RNP; guanosine nucleotide binding protein alpha subunit, GNAL, a candidate autoantigen in autoimmune hypophysitis; and Ku 70/80, respectively, ***Fig. 1, middle panel***). HC06 and HC30 each had high MFI anti-thyroperoxidase (TPO) that exceeded the 5 SD cutoff if excluded from calculating the average TPO MFI using the other 29 HC samples. Both samples were therefore considered “positive” in our analysis, but we included them in calculating the 5 SD cutoff on the COVID samples. In striking contrast, 25 of 51 (49%) hospitalized patients with COVID-19 had autoantibodies recognizing at least one autoantigen (***Fig. 1, top panel***). Using a stringent 5 SD cut-off, serum antibodies from eleven COVID-19 patients identified a single antigen, thirteen recognized 2-3 antigens, and one subject (Subject UP40) recognized nine different autoantigens. Ribosomal P proteins (P0, P1, and P2) were most prominently targeted in patients (10 of 50 patients, 20%), but were not found in any of the HC. Similar results were observed in 48 Kaiser subjects analyzed using an earlier-generation 26-plex autoantigen microarray, identifying overlapping RNA-containing autoantigen complexes including RPP14 Th/To, the Ro/La particle, the U1-small nuclear ribonuclear protein (U1-snRNP), thyroid antigens, and chromatin proteins as targets in hospitalized COVID-19 patients, but in none of the HC (***Supplementary Fig. 4***).

Rare antigens seen in patients with autoimmune myositis (MDA5, Mi-2, and tRNA synthetases such as PL-7 and Jo-1), and candidate autoantigens in autoimmune myocarditis (troponin and MYH6, ***Fig. 2a***), were observed in individual patients, as were rare SSc autoantigens (Th/To (RPP25), fibrillarin, and the U11/U12 snRNP, ***Fig. 2b***). A subset of autoantibodies (e.g., antibodies that bind the complement inhibitor C1q, thrombosis-associated antibodies that target beta 2 glycoprotein 1 ( 2- β GP1), and vasculitis-associated antigens such as bactericidal permeability inducing protein (BPI)) that have been implicated in pathogenic inflammation in target organs, were also found in individual patients (***Fig. 2c***)^4-6 14-16^. Relatively common autoantigens such as Scl-70, CENP A/B, and Sm/RNP were infrequent. Thyroid autoantibodies were also commonly observed (12/147 subjects across our entire study, 8.2%, using cutoffs of 3,000 MFI and 5 SD above HC). Thyroid dysfunction, which is relatively common in the general population, has been reported in COVID-19 patients^4,5^. In all cases where samples from more than one time point were available, anti-TPO and anti-thyroglobulin (TG) were already present at high MFI levels in the baseline sample. Taken together, these findings reveal that hospitalized patients with COVID-19 produce an increased frequency of autoantibodies, but that there is substantial inter-individual variation in which autoantigens are targeted.

**Fig. 2:**
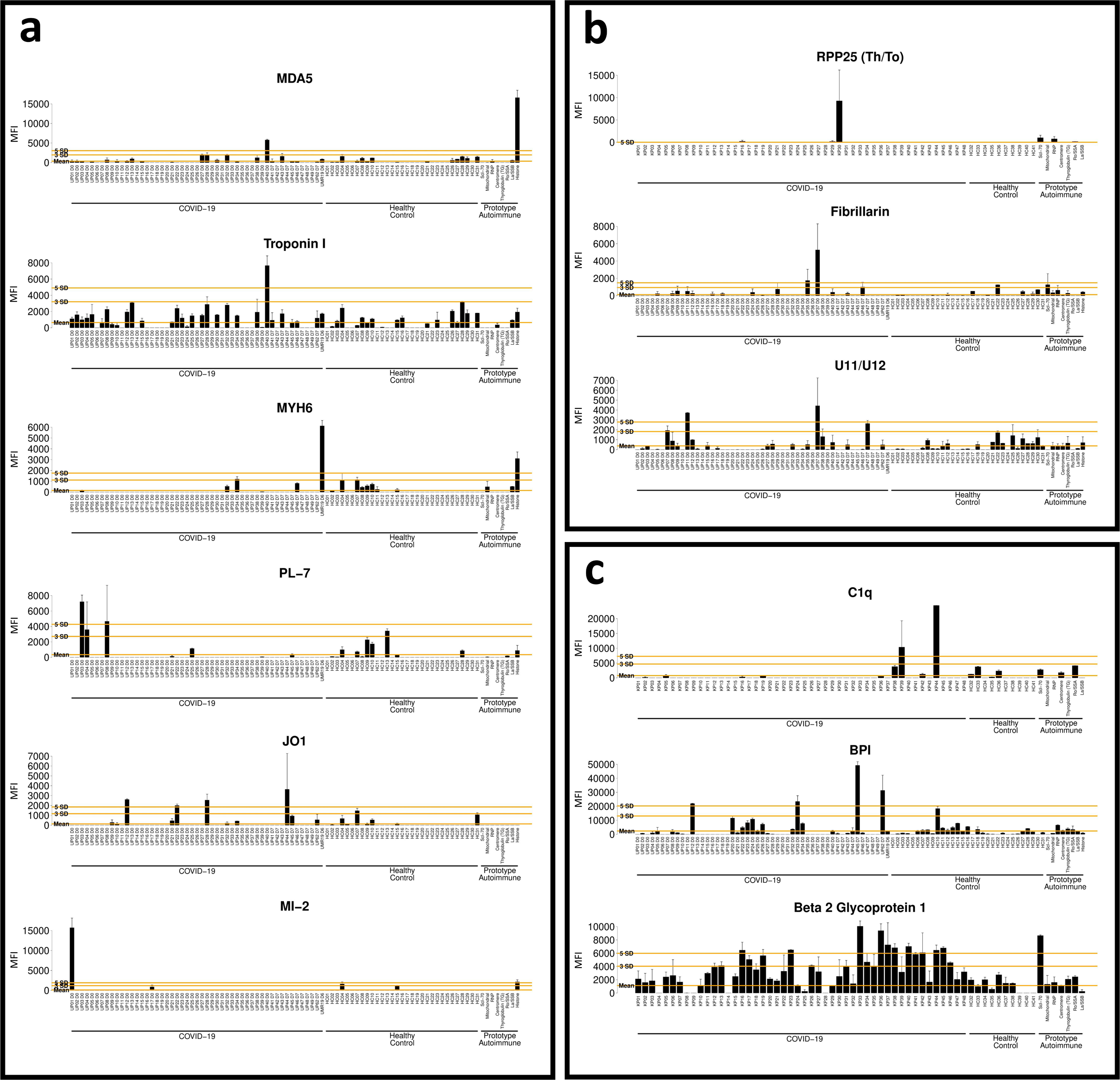
Serum autoantibodies in COVID-19 patients recognize antigens targeted in rare connective tissue diseases, and antigens associated with pathogenicity. Bar plots of twelve antigens corresponding to Figure 1. **a.** Antigens associated with autoimmune myositis and myocarditis (MDA5, troponin 1, MYH6 (alpha-myosin), PL-7, Jo-1, and Mi-2). **b.** RNA-containing antigens associated with systemic sclerosis (RPP25 Th/To, Fibrillarin, and U11/U12). **c.** Antigens that may be pathogenic (C1q and β2GP1) and associated with vasculitis (BPI). MFI are shown on the y-axis. Subjects are shown on x-axis (COVID-19 patients, HC, and Prototype Autoimmune). Average MFI for HC, 3 SD above the average MFI for HC, and 5 SD above the average MFI for HC are shown with orange lines. Error bars represent one standard deviation of the MFI for sample replicates.

### Secreted proteins are common autoantigens in hospitalized COVID-19 patients

In a pair of elegant studies, Bastard^5^ and Wang^17^ independently identified anti-cytokine antibodies (ACA) in patients with severe COVID-19. Both groups showed that a subset of ACA prevent binding of soluble factors to their cognate cell surface receptors and have been postulated to play a pathogenic role by thwarting protective immune responses to COVID-19. We created a 41-plex array comprising secreted proteins and cell surface receptors, modeled on arrays we and others have used previously to characterize “secretome” antibodies in immunodeficiency disorders^18, 19^, SLE^18^, and systemic sclerosis patients^20^ (***Supplementary Table 2***). We observed even more striking results with the secretome array, which revealed that serum antibodies in 41/51 (80%) of hospitalized COVID-19 patients recognized at least one secreted or cell surface autoantigen (***Fig. 1, upper right half of panel***), while only 2/31 (6%) HC subjects recognized a single antigen (interferon-gamma, IFN-γ in one and CD74 in the other, ***Fig. 1, middle right half of panel***). Interestingly, the IFN- + HC γ subject (HC27) also had serum antibodies specific for Sm (a subunit of the U1-snRNP, using 5 SD cutoff) and for both Ro60 and La (using a 3 SD cutoff), suggesting this “healthy” subject is in preclinical evolution toward developing SLE, a disease in which we have previously described multiple different ACA including anti-IFN-α and anti-B cell activating factor (BAFF).

Interferons, particularly the Type I interferon IFN-α were targeted in multiple COVID patients at frequencies (n=23 across all interferons, 45%) higher than recently published findings from other groups^17, 21^. Serum from five subjects (UP11, UP38, UP41, UP42, and UP46) recognized two or more interferons. In some COVID-19 patients, MFI values were comparable to or even exceeded those observed in previously characterized prototype patients with autoimmune polyendocrine syndrome type 1 (APS-1), pulmonary alveolar proteinosis (PAP), and atypical mycobacterial infections (AMI) (see ***Fig. 3d***).

**Fig. 3:**
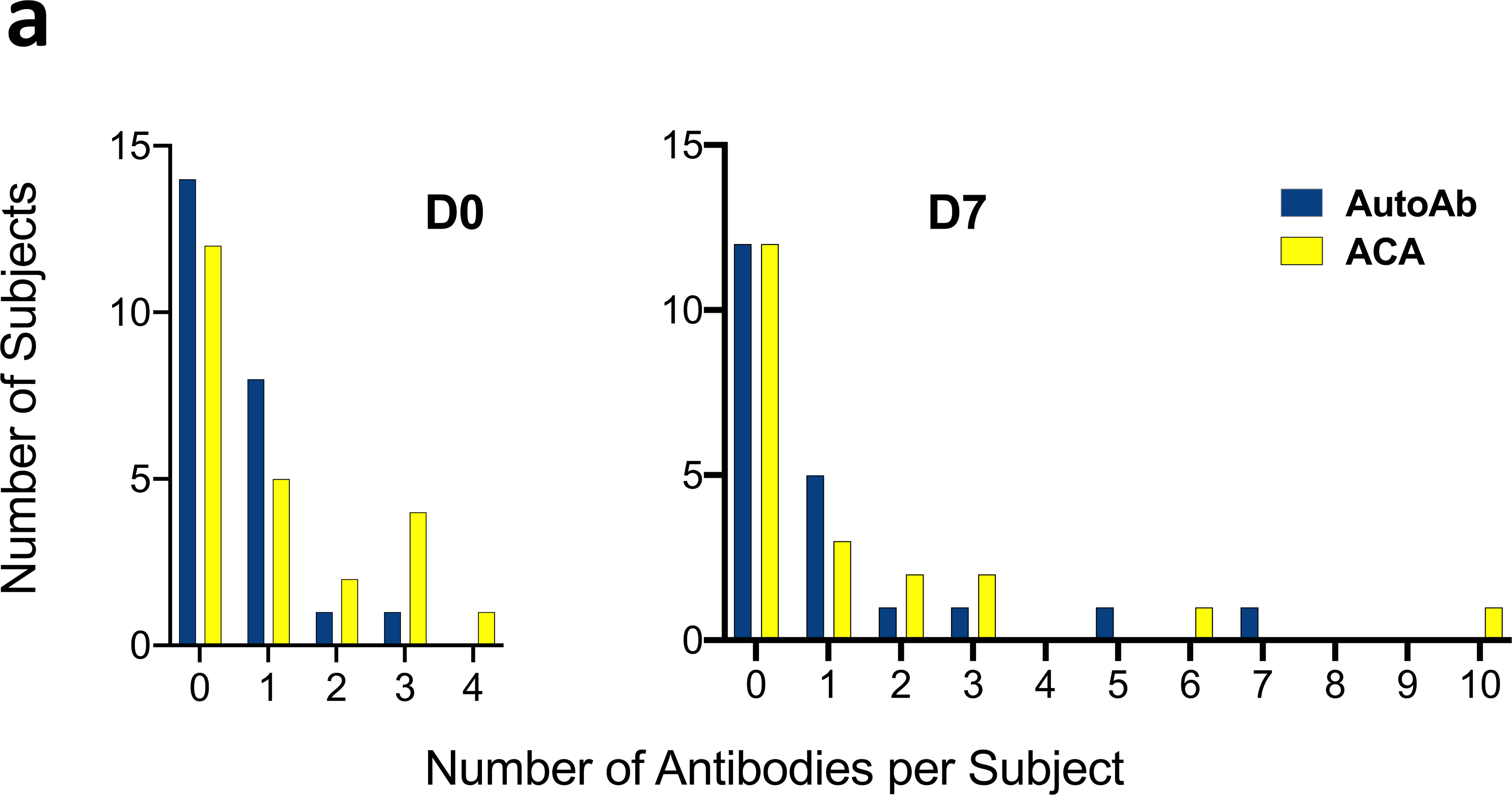
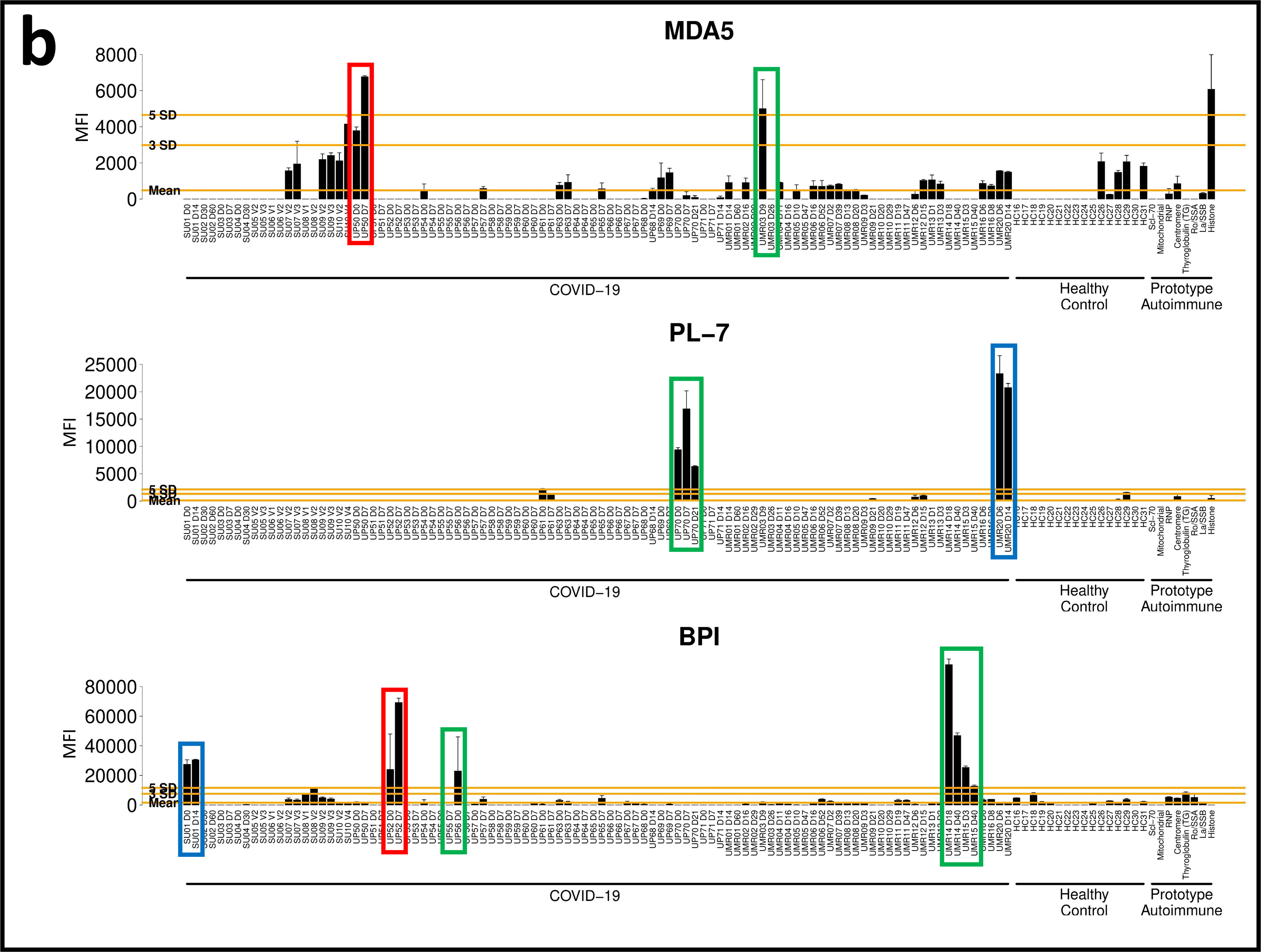
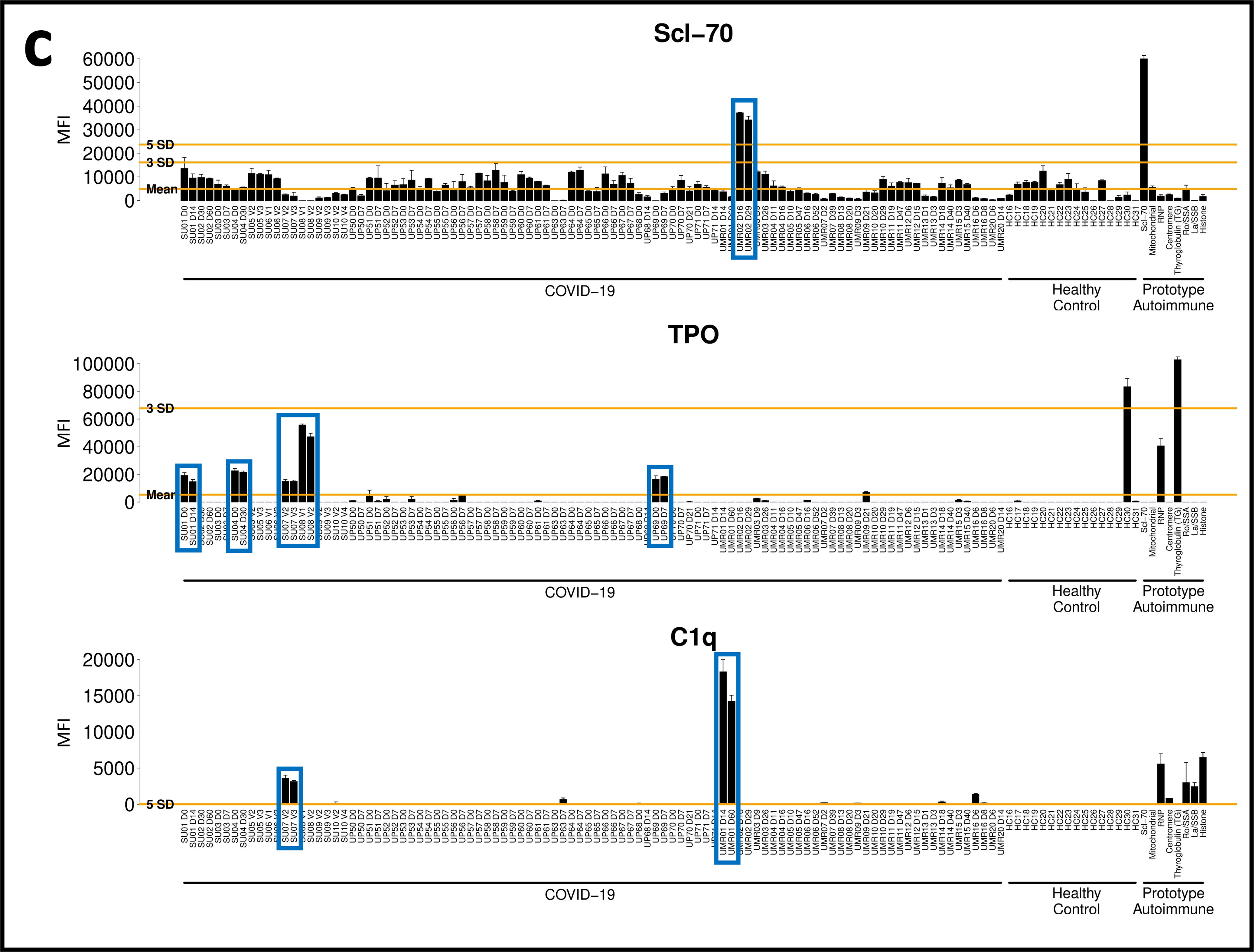
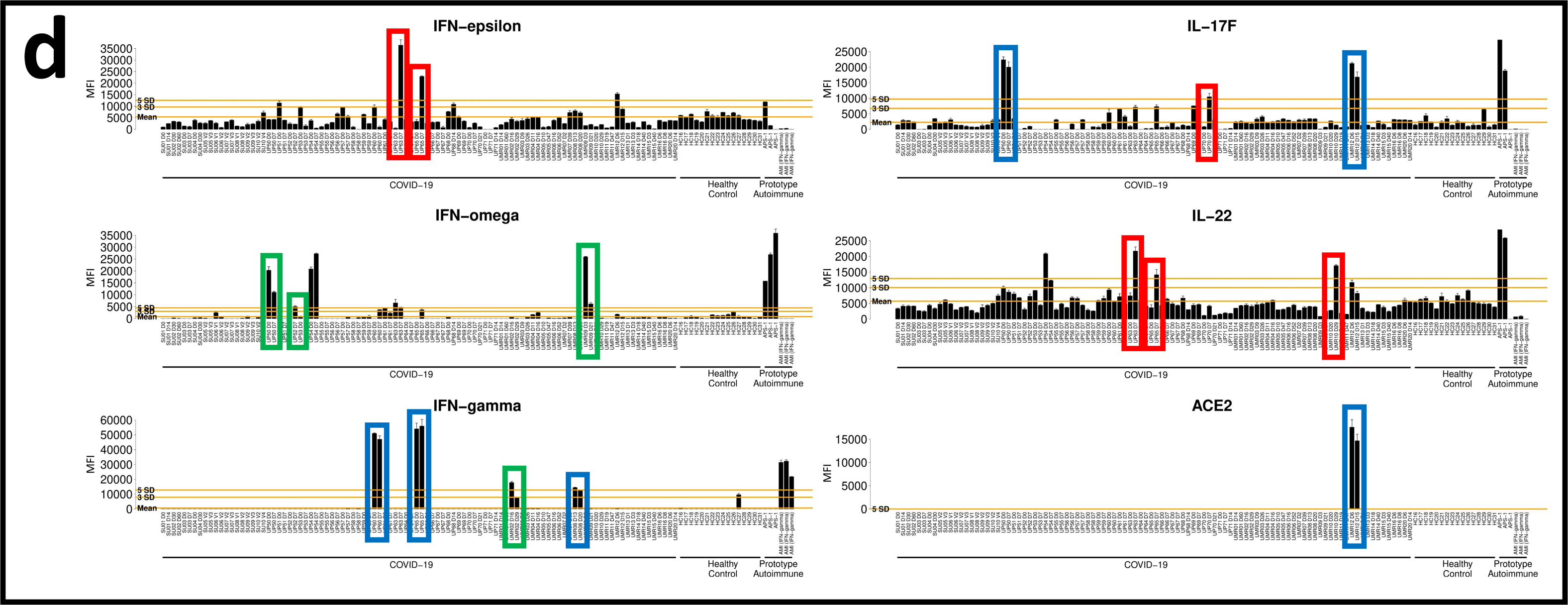
Autoantibodies are triggered by SARS-CoV-2 infection. **a.** Autoantibody (AutoAb, blue) and anti-cytokine antibody (ACA, yellow) counts are shown at Day 0 (n=24 patients, left) and Day 7 (n=21 patients, right). Counts were based on antibodies that were present at levels at or exceeding 5 SD above the average MFI for healthy control samples. **b.** Examples of transient or fluctuating autoantibodies against MDA5, the tRNA synthetase PL-7, and the vasculitis antigen BPI are shown. **c.** Examples of antigens (Scl-70, TPO, and C1q) that are likely to have been present at the time of infection and are unaffected by SARS-CoV-2 infection, are shown. **d.** Examples of ACA that are inducible (e.g., IFN-ε), or are present at baseline with little change over time (e.g., IFN- γ), are shown. Additional examples (IL-17, IL-22, and ACE-2) are also included. MFI are shown on the y-axis. Subjects are shown on the x-axis (COVID-19 patients, HC, and Prototype Autoimmune). Red boxes signify autoantibodies that increase over time. Green boxes signify autoantibodies that decrease over time or are transient (e.g., anti-PL-7, subject UP70). Blue boxes signify antibodies that are elevated at baseline and remain relatively unchanged over time. Average MFI for HC, 3 SD above average MFI for HC, and 5 SD above the average MFI for HC are shown with orange lines. Error bars represent one standard deviation of the MFI for sample replicates.

Many interleukins were also prominently identified as autoantibody targets in this screen (e.g., interleukins −1, −6, −10, −15, −17A, −22 and −31), as were cytokines with well-characterized functions such as leukemia inhibitory factor (LIF), the chemotactic chemokine macrophage inflammatory protein-1 alpha (MIP-1α), and vascular endothelial growth factor-B (VEGF-B). Several striking reactivities were observed in individual COVID-19 patients, including IL-12p70 (Subject UP47); the SARS-CoV-2 receptor angiotensin converting enzyme-2 (ACE-2, Subject UMR19); granulocyte macrophage colony stimulating factor which is the causative autoantibody target in PAP (GM-CSF, Subject UP25); oncostatin-M (OSM, Subject UP40); and soluble receptor activator of nuclear factor kappa B (sRANK-ligand, Subject UP19). Subject UP17 was being treated with a tumor necrosis factor-alpha (TNF-α) inhibitor at the time of SARS-CoV-2 infection, explaining the high MFI reactivity to TNF (***Fig. 1***). MFI for all antigens except IL-12p70 were very high (>10,000) in individual patients. Autoantibodies against all interleukins, cytokines and ACE-2 identified in the initial screen were also observed using a 5 SD cutoff in a second COVID-19 cohort (n=98 longitudinal samples from 48 different patients, see ***Fig. 3*, *Supplementary Figs. 5 and 6***), with few exceptions (e.g., IL-1 α, although IL-1 β was targeted using a 3 SD cutoff; IL-31, which met a 3 SD cutoff; and GM-CSF).

### A subset of autoantibodies is triggered by SARS-CoV-2 infection

To determine if autoantibodies and anti-cytokine antibodies were generated *de novo* (versus existed prior to infection), we analyzed 48 hospitalized COVID-19 patients (Stanford University, University of Pennsylvania, and Marburg University) in whom samples were available at two or more different time points. Twenty-four patients had an available sample from the day of hospitalization or t (day 0). The interval between collection of the second sample ranged from 2-58 days (mean interval = 15.8 days). Two subjects (UP70 and UP71) also had a third sample drawn 14-21 days post ICU admission. To reduce batch effects, all samples at all time points were analyzed on the 53-plex COVID-19 Autoantigen Array in the same instrument run (***Supplementary Fig. 5 top panel***) together with HC (***Supplementary Fig. 5, middle panel,*** n=16) and serum samples from prototype autoimmune diseases served as positive controls (***Supplementary Fig. 5, bottom panel***, n=8).

As with the unpaired samples described in ***Fig. 1***, autoantibodies from patients with paired samples had high MFIs in individual patients. Some patients were again identified whose serum recognized a large number of autoantigens (***Supplementary Figs. 5 and 6***). Twenty-five (52%) of hospitalized COVID-19 patients had autoantibodies against at least one autoantigen. Serum autoantibodies recognized two or more antigens (range 2-7 antigens) in seven patients (15%) (***Fig. 3a, and Supplementary Fig. 6***). Longitudinal analysis identified prominent increases in autoantibodies at the second available time point (***Fig. 3b,*** red boxes). In 9 individual patients (19%), autoantibody measurements were above the average for HC at the earliest available time point and MFI increased by at least 50%, exceeding the 5 SD and 3,000 MFI cutoff at the later time point (***Fig. 3b***, e.g., MDA5, subject UP50; BPI, subject UP52; ***Supplementary Fig. 6***). Some autoantibodies were at or below the average for HC at the first time point and increased over time (e.g., histones and histone H3, subject UP65; and β 2GP1, subjects UP65 and UP52), suggesting these autoantibodies were directly triggered by SARS-CoV-2 infection. Others were already elevated at the first time point and did not have large increases in MFI over time (n=22, 45%, blue boxes) (***Fig. 3c and Supplementary Fig. 6***). In a small number of cases, autoantibody MFI levels decreased below the SD and MFI cutoffs over time (n=5, 10%, green boxes), suggesting that their development might be transient (e.g., PL-7, subject UP70, ***Fig. 3b***). Anti-TPO and anti-Scl-70 (***Fig. 3c***, blue boxes) remained elevated at high levels in all seropositive subjects regardless of the time of measurement, suggesting that these autoantibodies were already present at hospitalization and likely represent preclinical (asymptomatic), unreported, or undiagnosed autoimmunity.

To further evaluate the potential evolution of autoantibodies, we performed ANA testing on 21 individuals with paired samples. Eight individuals (38%) had positive or weak positive ANA reactivity. Among these 8 individuals, ANAs were present at both time points in three, changed in intensity of staining in two, and were positive at only one of the two time points in the final three (***Supplementary Fig. 1b***). Taken together, these data indicate that autoantibody levels change over time in individual COVID-19 patients, consistent with their production and, in some cases, transience during acute illness.

We next examined whether IgG ACA are triggered by SARS-CoV-2 infection. Paired samples from the same 48 subjects described above were used to probe the 41-plex cytokine array, again in a single, batched run. As observed with unpaired samples (***Fig. 1***), 28 of 48 (58%) of COVID-19 patients had at least one ACA (***Supplementary Fig. 6***). Of these twenty-eight, sera from fifteen patients recognized one cytokine, five recognized two cytokines, and eight recognized three or more cytokines (range 3-12 antigens). Interferons, IL-17, and RANK-L were the most common targets, and interferons, IL-17, and IL-22 were new targets in some patients (***Fig. 3d***). In addition to Subject UMR19 (***Fig. 1***), a second patient with high MFI ACE-2 autoantibodies was also identified (Subject UMR12, ***Fig. 3d***). Increased MFI was observed for one or more autoantibody at later time points in 12 patients (24%). Several were present at MFI levels near or below the average for HC at baseline and were induced to high MFI levels at the second time point (e.g., anti-IFN-α, subject UMR07; anti-IFN-ε, subjects UP63 and UP65; and anti-IL-22, subjects UP54, UP63, UP65, and UMR10, ***Fig. 3d***). In many patients, ACA MFI levels were significantly elevated at the first time point and decreased at the later time point, suggesting that some ACA were pre-existing and/or developed transiently following SARS-CoV-2 infection. Subject SU09 had very high MFI levels of anti-TNFα at both time points, attributed to anti-TNF therapy. We conclude that antibodies against cytokines, chemokines, growth factors, and receptors are common in hospitalized COVID-19 patients. Many are triggered in response to SARS-CoV-2 infection, even at later time points distant from the time of infection (e.g., anti-IL-22, subject UMR10, day 29, ***Fig. 3d***).

To further evaluate the change in autoantibodies and ACA over time, we performed a targeted analysis of 21 of the 48 patients who had paired autoantibody and ACA data specifically at D0 and D7 of hospitalization (***Supplementary Fig. 7***). Almost all patients (18/21, 86%) had demonstrable changes in the number of antibodies, defined at varying thresholds of sensitivity (>3 SD vs. 3-5 SD vs. >5 SD) between D0 and D7 (***Supplementary Fig. 7a***). When combining the number of autoantibodies or ACA (***Supplementary Fig. 7b***), there is a trend towards increased numbers both of autoantibodies and ACAs per subject over time. Higher numbers of individuals with more autoantibodies and ACAs at D7 compared to D0 at the 3-5 SD threshold were observed (***Supplementary Fig. 7b and 7c***), but the difference in medians between D0 and D7 was not statistically significant. Nevertheless, these data clearly show that there is ongoing evolution in both the numbers and levels of autoantibodies and ACAs with time in hospitalized COVID-19 patients.

### Broad anti-viral immune responses target internal viral proteins in hospitalized COVID- 19 patients

We have used protein arrays for epitope mapping and to measure antibody responses in influenza vaccines^22^ and in a nonhuman primate human immunodeficiency virus (HIV) vaccine study^23^. We used a similar approach here to characterize anti-viral responses following SARS-CoV-2 infection. We created a 28-plex COVID-19 viral array that included structural and surface proteins from SARS-CoV-2 as well as eight commercially available recombinant nonstructural proteins localized to the interior of the virus (***Supplementary Table 3***). As an initial validation, we compared array-based detection and measurement using a clinical-grade ELISA (R=0.81, Spearman’s, p<0.0001 for anti-SARS-CoV-2 nucleocapsid; R=0.60, Spearman’s, p<0.0001 for anti-SARS-CoV-2 RBD, ***Supplementary Fig. 8a and 8b, respectively***). By studying the anti-viral antibody (AVA) response, we hoped to understand if certain viral antigens might correlate with the development of autoimmune responses. We hypothesized that poorly controlled SARS-CoV-2 infection leads to the development of serum antibodies that recognize not just structural proteins such as the SARS-CoV-2 spike protein, but also nonstructural proteins, and that a subset of these viral proteins might correlate with the development of autoimmunity. Proteins from related coronaviruses were also included to explore whether pre-existing antibodies to seasonal coronaviruses might correlate negatively or positively with disease severity, and with autoimmunity.

*Fig. 4* depicts a heatmap representation of IgG reactivity based on MFI (Fig. 4a***, left panel***) and calculation of SD above average MFI for HC (Fig. 4b***, right panel***). As expected, nearly all patients had broad immune responses to viral structural proteins (first seven antigens on left, Fig. 4a and b). Twelve patients had low MFI levels at the earliest time point (almost all were day 0, defined as collection within the first 24-72 hours of hospitalization but developed high MFI IgG SARS-CoV-2 antibodies when tested at later time points, consistent with previously published findings in the setting of acute illness^24^ (Fig. 4a). Other subjects (e.g., subject UP50) already had broad AVA responses at day 0, suggesting they had been infected for a significant period of time prior to hospital admission.

**Fig. 4:**
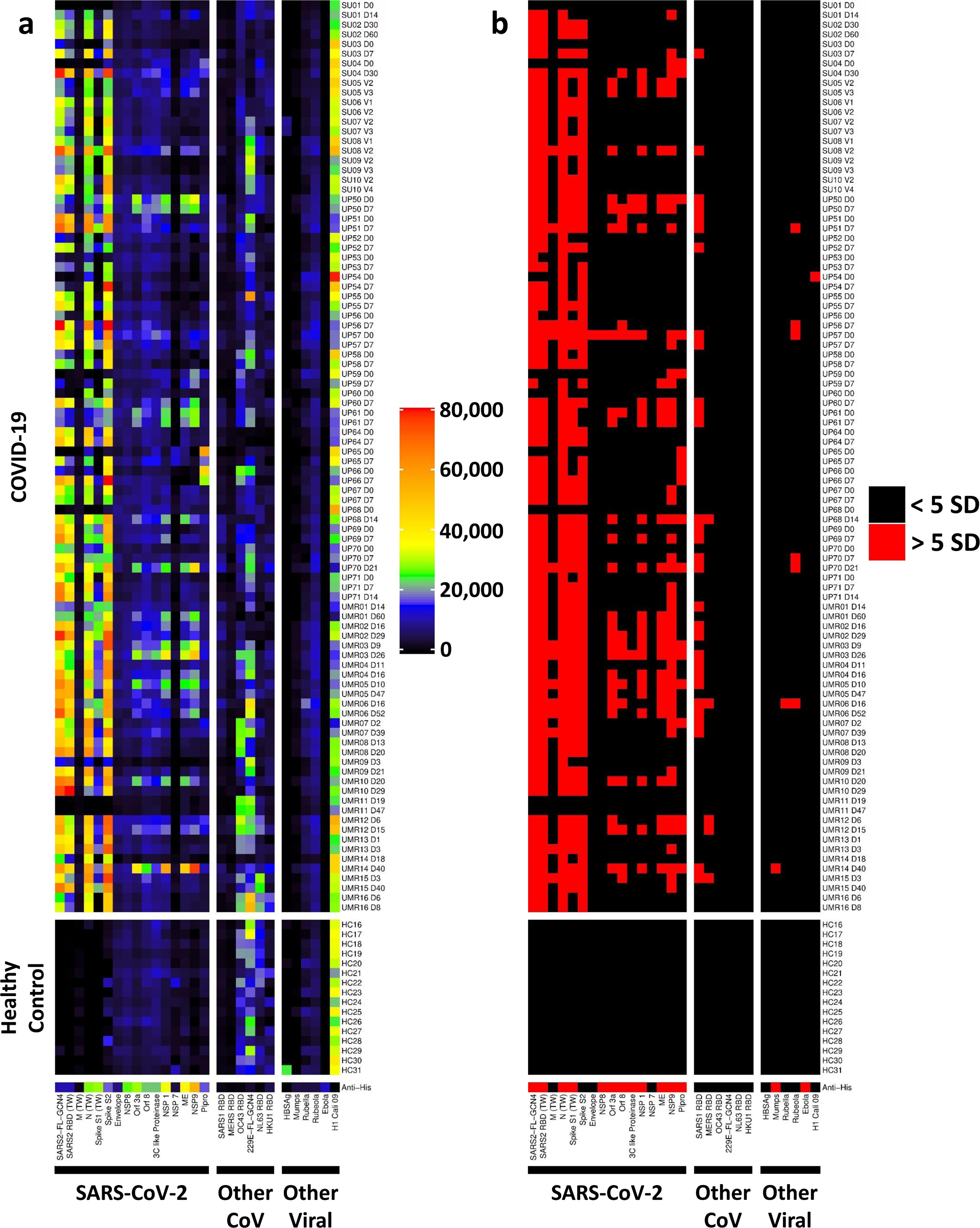
Measurement of anti-viral IgG responses using a COVID-19 viral array. **a.** Heatmap depicting IgG antibodies using a 28-plex bead-based protein array. Viral protein antigens are grouped based on sixteen proteins from SARS-CoV-2 (left panel), other coronaviruses (middle panel) and other viruses (right panel), labelled on the x-axis. Most recombinant viral proteins were engineered to include a 6X-His-tag, which was used to validate conjugation to beads using an anti-epitope monoclonal antibody (bottom of panel). The same COVID-19 patients from Figure 3 (see Supplementary Figure 6) were analyzed (top panel, n=94 longitudinal COVID-19 samples, including paired samples from 44 subjects and 2 subjects who had 3 available timepoints each, subjects UP70 and UP71). HC (n=16, middle panel). Two patient sample pairs (UP63 and UMR20) and were excluded from analysis due to technical failure on the viral array assay. Colors correspond to the MFI values shown at right. **b.** Heatmap depicting statistically significant anti-viral IgG responses. Colors indicate IgG antibodies whose MFI measurements are >5 SD (red) or <5 SD (black) above the average MFI for HC samples collected prior to the COVID-19 pandemic.

IgG antibody levels against non-structural SARS-CoV-2 proteins were significantly elevated in a majority of patients (n=35, 73%), particularly papain-like protease (PLPro, n=13 patients), open reading frame proteins (Orf 8, n=14 patients and Orf 3a, n=18 patients); and nonstructural proteins (NSP 1, n=20 patients, and NSP 9 n=31 patients, but not NSP 7 (n=0), NSP8 (n=1) or 3C-like protease (n=3)). The number of targeted non-structural proteins increased over time in 20 of 49 (40%) patients when compared with the earliest available time point, and were absent (n=14), did not change (n=7), or decreased (n=8) in the remaining patients. Of the eight patients who showed a decrease in the number of targeted non-structural SARS-CoV-2 antigens over time, all but one decreased by a single antigen, and three were patients had samples collected at an interval of 37 days, making it likely that the immune responses were transient. A majority of patients had linked antibody responses in which multiple non-structural antigens were targeted in the same subject. In rare patients (e.g., subject UP65, see SARS-CoV-2 protein PLpro, Fig. 4a and 4b), the initial immune response was focused on an internal protein (or was pre-existing) and later evolved to target spike and other SARS-CoV-2 surface or structural proteins. We conclude that antibody responses in hospitalized COVID-19 patients are not limited to structural proteins, that linked responses to multiple non-structural proteins are observed over time, and that NSP9 is the most commonly recognized internal SARS-CoV-2 protein of those tested on the array.

### New-onset IgG autoantibodies are temporally associated with anti-SARS-CoV-2 IgG responses

We next identified a subgroup of patients (n=12) whose anti-SARS-CoV-2 antibody responses suggested that they had been infected at a time point that was proximate to hospitalization and capture of the first sample. Selection criteria for patients who were early in their anti-viral responses included (i.) the first available sample was within three days of hospitalization; (ii.) anti-spike S1 IgG levels were <5,000 MFI at baseline; (iii.) anti-RBD IgG levels were <20,000 MFI at baseline; and (iv.) at least a 2-fold increase in MFI for IgG against both S1 and RBD was observed at the next available timepoint. We then studied these patients to further determine if new IgG autoantibodies appeared at the second time point, providing evidence that SARS-CoV-2 directly triggers development of autoantibodies.

We compared IgG reactivities at both time points for all twelve subjects who met the above criteria on COVID-19 autoantigen arrays (Fig. 5a***, left panel***) and cytokine arrays (Fig. 5b***, middle panel***) with anti-viral responses using the virus array (Fig. 5c***, right panel***). Four of twelve patients were found to have at least one newly induced autoantibody at the later time point (white boxes). Two of these four patients had two or more new autoantibodies (Subjects UP52, n=5 antigens; and subject UP65, n=10 antigens). β2GP1, histones, and the 54 kD component of the myositis autoantigen signal recognition particle (SRP 54) were the most common antigens identified (n=2 subjects each). Given the small sample size, no clear correlations were identified between individual autoantibodies and an IgG response to a specific viral protein (Fig. 5d ***and Supplementary*** Fig. 9).

**Fig. 5:**
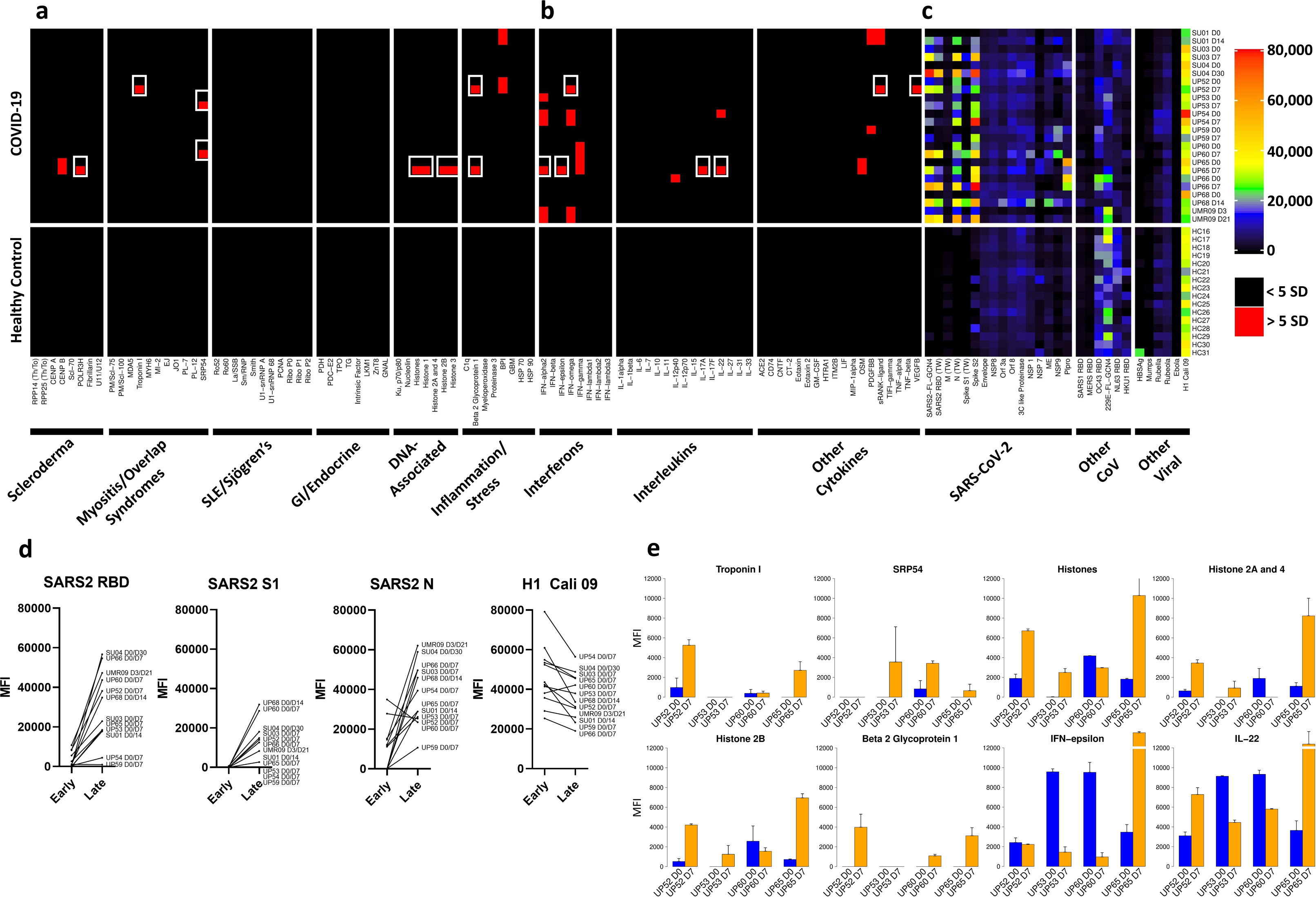
New-onset autoantibodies correlate with anti-SARS-CoV-2 IgG responses over time in recently infected patients who developed COVID-19. Twelve patients were identified who had low or absent anti-SARS-CoV-2 RBD or spike S1 protein responses at baseline and who went on to develop high MFI IgG SARS-CoV-2 antibodies at the next available time point. Autoantigen data (**a**) and ACA data (**b**) from Supplementary Figure 6 (using 5 SD and MFI >3,000 cutoffs) for these 12 patients and HC has been combined with anti-viral heatmap data from Figure 4 (**c**). Multiple new autoantibodies are depicted with white boxes. Antigens are shown on the x-axis. Patients and HC are shown on the y-axis. Colors for viral IgG levels correspond to the MFI values shown at far right. **d.** Line graph comparing MFI for IgG antibodies against four viral proteins at Early (D0) and Late (D7) time points for the same 12 patients in Panels A-C. **e.** Bar graph comparing MFI for IgG autoantibodies against eight autoantigens or cytokines at Early (D0) and Late (D7) time points for the same 12 patients in Panels A-D. Error bars represent one standard deviation of the MFI for sample replicates.

Finally, we correlated autoantibodies and ACA with anti-viral IgG responses using array data from the cohort described in Fig. 3, focusing on Penn and Marburg samples which had been collected at time points as proximate to the day of hospitalization as possible. We compared patients who had one or more autoantibody (n=15first time point, n=13 second time point) with patients who had no autoantibodies (n=21 first time point, n=23 second time point). Anti-SARS-CoV-1 RBD correlated positively with the autoantibody positive group (p=0.002 at the second time point using Wilcoxon rank sum test, p=0.044 using Bonferroni correction for each time point). NSP1 (p=0.03 at second time point, p=0.08 at first time point) and ME (p=0.04 at first time point; p=0.06 at second time point) trended positively when correlating with autoantibodies but were not statistically significant when correcting for multiple hypotheses. An identical analysis was performed on ACA+ vs. ACA- patients, showing no correlations with IgG responses to any viral proteins, including influenza, SARS-CoV-1, and seasonal coronaviruses (OC43 RBD, 229E-FL-GCN4, NL63 RBD, and HKU1 RBD).

## Discussion

We have used a multiplexed, bead-based platform to identify circulating antibodies in hospitalized patients with COVID-19 and have generated integrated results from three different protein microarrays to discover COVID-19 associated autoantigens and link them to anti-viral responses. Our studies have led to several important findings that provide further insights into COVID-19 pathogenesis. First, we found that approximately half of hospitalized COVID-19 patients develop serum autoantibodies against one or more antigens on our array even though only a quarter of all patients are ANA+. Increased levels of autoantibodies are not simply a reflection of hypergammaglobulinemia because they are produced out of proportion to total IgG serum concentration. In most individuals, only a small number of autoantigens are targeted, which is more consistent with a sporadic loss of self-tolerance than a global increase in autoantibody production. Second, the autoantibodies we discovered are found in relatively rare connective tissue diseases that are not typically measured in clinical labs, and some are predicted to be pathogenic. Third, a surprisingly large number of ACA were identified, far more than just the interferon autoantibodies described recently^21^. Fourth, antibodies recognizing nonstructural SARS-CoV-2 proteins were identified that correlate positively with autoantibodies. Finally, and perhaps most importantly, some autoantibodies are newly triggered by SARS-CoV-2 infection, suggesting that severe COVID-19 can break tolerance to self.

Approximately 60-80% of all hospitalized COVID-19 patients in our study had at least one ACA, with a greater number of different ACA specificities generated in individual patients than observed for traditional autoantigens. Two recent studies demonstrated that IFN-α and IFN-ω- blocking ACA are found in patients with severe COVID-19^21, 25^. Anti-IFN antibodies with blocking activity were absent in all 663 tested patients with mild COVID-19, strongly linking the presence of anti-IFN to disease pathogenesis and severity^21^. Another study reported that type I interferon (IFN) deficiency could be a hallmark of severe COVID-19^26^, while other investigators pointed towards an untuned antiviral immune response due to delayed type I/III interferon expression^27^. Bastard identified blocking ACA for additional cytokines including IL-6, IL-22, and IL-12p70^21^. ACA without blocking activity still may be biologically important, for example by potentiating receptor binding or prolonging cytokine half-life^28, 29^. In another recent study, Wang identified autoantibodies against additional secreted and tissue-associated proteins in COVID-19 patients^17^, some of which were pathogenic when tested in animal models of SARS-CoV-2 infection. Pre-existing IFN-α autoantibodies were recently identified in 4/10 (40%) SLE patients from NIH’s SLE cohort who later became infected with SARS-CoV-2^32^. We have identified ACA in SLE (including anti-BAFF blocking antibodies and anti-IFN α)^18^, systemic sclerosis^30^, and a variety of immunodeficiency disorders^18,19,31^, suggesting that ACA are probably more common than previously appreciated in immune-mediated diseases. Taken together, these earlier studies are consistent with the notion that pre-existing ACA are pathogenic and may place such individuals at increased risk of developing severe COVID-19. What is different about our work from these earlier studies is that we show a change in ACA levels and in the numbers of ACAs over time in many hospitalized individuals with acute COVID-19. Our findings suggest that ACA may also form in response to viral infection or as a consequence of an inflammatory immune response in which high levels of cytokines are generated.

In addition to ACAs modulating the immune response and potentially causing more destructive inflammation, autoantibodies have the potential to contribute in a number of other ways to COVID-19 pathogenesis. Several autoantigens we discovered are naturally complexed with a structural RNA molecule which could serve as a ligand for nucleic acid sensors such as Toll Like Receptors (e.g., TLR7, TLR3) in host cells. RNA or DNA released from dying cells could also form immune complexes with viral or self-antigens that can promote autoantibody production. A subset of array-identified autoantigens (e.g., MDA5) are encoded by interferon-inducible genes and would be predicted to be transcribed in response to SARS-CoV-2 infection. Indeed, the acute phase of severe SARS-CoV-2 infection can be accompanied by marked tissue inflammation, cytokine storm (including secretion of interferons), upregulation of interferon signaling pathways, and expression of ACE-2 in vascular endothelium. Although not yet explored for COVID-19-associated autoantibodies or ACA, IgG antibodies that bind SARS-CoV-2 proteins are often IgG1 and have afucosylated glycans. These properties enhance immunoglobulin interactions with the activating Fcγ receptor FcγRIIIa, potentially leading to increased production of inflammatory cytokines such as IL-6 and TNF^33^.

We postulate that a subset of the autoantibodies we have identified contribute to the formation of inflammatory immune complexes *in situ*, particularly at endothelial surfaces. For example, neutrophil extracellular traps (NETs) have been implicated in COVID-19 patients with vasculitis.^34^ Antineutrophil Cytoplasmic Antibody (ANCA) associated vasculitis (AAV) has been strongly associated with neutrophil activation and generation of proinflammatory NETs containing nucleic acids, histones, and inflammatory peptides^35^. While we did not observe elevated levels of MPO or PR3, we identified high MFI anti-BPI antibodies in 6% of COVID-19 patients. The detection of autoantibodies to BPI and core as well as linker histones raises the possibility that NETs contribute to the generation of autoantibodies in severe COVID-19, a possibility that is in line with the neutrophilia that accompanies severe acute disease^36^. Disseminated microvascular coagulopathy and microvascular injury in lung and skin from COVID-19 patients correlate with fibrin deposition and thrombus formation^37^. SARS-CoV-2 membrane proteins including the spike protein (but not SARS-CoV-2 RNA) colocalize with activated complement in ACE-2+ microvascular endothelia of COVID-19 lung tissue and normal-appearing skin^37, 38^. Magro and colleagues hypothesize that spike protein on the surface of circulating pseudovirions binds to endothelial ACE-2 (whose gene is interferon-inducible), providing a nidus for activation of complement and formation of microthrombi. Anti-C1q (an SLE autoantigen), anti- β 2GP1 (which is thrombogenic), anti-BPI, and anti-ACE-2 (if non-blocking)^39^ that were discovered in our screen would be predicted to exacerbate these pathogenic processes^40^.

Severe infection may also result in an “all-hands-on-deck” immune response that results in loss of tolerance due to the presence of pro-inflammatory mediators that may lessen the requirement for T cell help. Some patients with severe acute COVID-19 appear to mount extrafollicular B cell responses that are characterized by expanded B cells and plasmablasts, loss of germinal centers, and loss of expression of Bcl-6^41, 42^. Antibody repertoires analyzed from hospitalized COVID-19 patients during acute disease include massive clones with low levels of somatic mutation (SHM)^43^,^44^ and elongated CDR3 sequences which can be associated with polyreactivity^45^ and are reminiscent of immune responses seen in acute Ebola^46^ and salmonella infection^47^. It has been suggested that these responses resemble SLE flares in which autoreactive B cells are also activated via an extrafollicular, TLR7-dependent pathway^8,41,48^. Although SARS-CoV-2 genomic RNA could itself serve as a costimulatory TLR7 ligand, many of the autoantigens we have identified also bind to structural RNAs such as the U1-snRNA (found in Sm/RNP complexes), 7S RNA (a component of SRP), and tRNAs (e.g., Jo-1, PL-7, and PL-12) which might activate dendritic cells in a TLR7- dependent manner^49, 50^.

One of the most important unanswered questions raised by our studies is why specific molecules are targeted in hospitalized COVID-19 patients. For newly triggered ACA, the most likely explanation is that they arise as a consequence of severe disease along with high levels of viremia, tissue injury, and elevated local levels of pro-inflammatory cytokines and chemokines. However, it is also possible that the presence of ACA could affect the regulation of self-reactive lymphocytes by altering the half-lives of the receptor interactions of the target molecules. For traditional autoantigens, one possibility is that viral proteins or the SARS-CoV-2 RNA genome and self-molecules physically interact, and that the initial immune response to the viral protein in a highly inflammatory microenvironment expands to include self-proteins through linked recognition and intermolecular epitope spreading. Another possibility is molecular mimicry in which one or more viral proteins or epitopes cross reacts with self-proteins leading to loss of tolerance and development of autoimmunity^51, 52^. Experiments to explore these mechanisms are ongoing.

The vast majority of studies on SARS-CoV-2 proteins have focused on viral structural proteins to develop efficient and accurate diagnostic assays, and to identify specific epitopes on surface proteins for development of vaccines and therapeutic monoclonal antibodies. These proteins are also the major focus of the immune response in most infected individuals. Here, we developed a multiplexed viral protein array that enables simultaneous measurement of antibody responses against 28 different proteins from 13 different viruses. We determined that non-structural proteins are recognized by antibodies in a large proportion of hospitalized COVID-19 patients, suggesting that B cell responses expand over time to involve additional viral molecules. IgG antibody levels against NSP1 and ME correlated positively with the presence of at least one autoantibody. We hypothesize that prolonged inability to eradicate and clear virus expands the adaptive immune response to target non-structural viral proteins, some of which might physically interact or cross-react with autoantigens in the context of an intense local or systemic inflammatory environment, exceeding a threshold for breaking tolerance to self. In contrast, patients who rapidly mount neutralizing antibody responses to the viral spike protein abort “intraviral epitope spreading” and may be less likely to develop autoantibodies. Why anti-SARS-CoV-1 RBD IgG responses associate with autoantibody positive patients is unclear. Future longitudinal studies are needed to determine whether broad B cell responses play any direct pathogenic role in patients with prolonged hospital courses or in patients with long-term sequelae of COVID-19 infection; to correlate anti-viral responses with ACA and autoantibodies over time using much larger COVID-19 cohorts including patients who are asymptomatic or have mild disease; and to explore whether specific SARS-CoV-2 proteins might cross-react with autoantigens discovered in our screens.

Many studies of hospitalized COVID-19 patients, including our study, suffer from important limitations. First, confounding variables exist including heterogeneous demographics, medications at hospitalization, individualized treatment approaches, and, in some cases, unknown history of pre-existing medical or autoimmune conditions. Second, “Day 0” is not day 0 of infection but instead refers to a time point most proximate to hospitalization. Our viral array results (Figs. 4 and 5) confirm that the time between initial infection and sample acquisition was heterogeneous, potentially confounding interpretation of autoantibody and ACA results. Third, not all antigens (e.g., lipids, hydrophobic proteins and carbohydrates) are compatible with our screening methodology, and as a result we have certainly missed some reactivities. Fourth, we did not include patients who were asymptomatic, had mild COVID-19, were vaccinated for SARS-CoV-2, had other severe viral illnesses, or were children. Finally, our analysis was limited to hospitalized patients during acute illness, with follow up times of days rather than months or years.

Although beyond the scope of these studies, our data generate many more questions that need to be addressed in the coming years – questions that can only be answered by generating large cohorts of prospectively enrolled subjects with new-onset viral syndromes, including patients with COVID-19, respiratory illnesses which resemble COVID-19, and subjects enrolled in COVID-19 vaccine trials. Are autoantibodies and ACAs specific to COVID-19, or is their presence shared more broadly in patients with influenza and other severe acute illnesses? Are autoantibodies and ACAs found in convalescent serum used to treat patients with severe COVID-19? Do any of these autoantibodies underly some of the signs and symptoms observed in “long COVID”, do they lead to classifiable autoimmune disease, and can they be used as predictive markers or to identify subsets of patients who would benefit from targeted immunotherapies?

Our studies have begun to quantify the impact of SARS-CoV-2 on autoimmunity, identifying which antigens and specific autoimmune diseases to surveil in patients who have been infected, and contributing to our mechanistic understanding of COVID-19 pathogenesis. These studies provide a starting point for large-scale epidemiology studies to determine the extent of autoimmunity that results from SARS-CoV-2 infection, and long-term impacts on the health care system and the economy. While the COVID-19 pandemic is leaving a wake of destruction as it progresses, it also provides an unprecedented opportunity to understand how exposure to a new virus could potentially break tolerance to self, potentially giving rise to autoimmunity and other chronic, immune-mediated, diseases.

## Methods

### Serum and plasma samples

#### Hospitalized COVID-19 patients

Serum or plasma samples were obtained following protocols approved by local institutional review boards (IRB) from 147 unique hospitalized subjects (n=99 unpaired; n=98 paired longitudinal samples from 48 distinct subjects). Samples were obtained from four centers in three distinct geographic areas: <u>Northern California</u> (Kaiser Permanente Health Care System, n=48 unpaired samples from hospitalized subjects, IRB# 55718) collected in March and April 2020; and Stanford Occupational Health Clinic, 20 paired samples from 10 unique hospitalized subjects IRB# 55689) collected between April and June 2020; <u>Philadelphia, Pennsylvania</u> (University of Pennsylvania, n=50 unpaired; and 44 paired samples from 21 unique hospitalized subjects, IRB # 808542) obtained between April and June 2020; and <u>Marburg, Germany</u> (Philipps University Marburg, 1 unpaired; and 34 paired samples from 17 unique hospitalized subjects collected between April and June 2020, IRB# 57/20). Clinical characteristics of the cohorts can be found in ***Supplementary Tables 4-6***.

#### Healthy Controls

Serum and plasma samples from anonymous healthy controls (HC, n=41) were obtained prior to the COVID-19 pandemic from Stanford Blood Bank and Stanford Hospital and Clinics.

### Bead-based antigen array content

We created three different custom, bead-based antigen arrays modelled on similar arrays that we previously used to study autoimmune and immunodeficiency disorders, and for characterizing vaccine responses^18,20,22,53-57^. Antigens were selected based on our published datasets; literature searches that have implicated specific antigens in COVID-19; potential for mechanistic contribution to COVID-19 pathogenesis; and compatibility with bead-based platforms. A complete list of all antigens, vendors, and catalogue numbers can be found in ***Supplementary Tables 1-3***.

The “COVID-19 Autoantigen Array” included 53 commercial protein antigens associated with CTDs (***Supplementary Table 1***). The “COVID-19 Cytokine Array” comprised 41 proteins including cytokines, chemokines, growth factors, acute phase proteins and cell surface proteins (***Supplementary Table 2***). Specific “secretome” proteins included a subset of molecules identified in previous large screens in patients with systemic lupus erythematosus (SLE) (59-plex screen)^18^, systemic sclerosis (scleroderma or SSc, 221-plex screen, manuscript in preparation)^30^, Autoimmune Polyendocrine Syndrome Type 1 (APS-1)^18^, Atypical Mycobacterial Infections (AMI)^18^, Immunodysregulation Polyendocrinopathy Enteropathy X-linked (IPEX)^19^, and more recently in COVID-19^9^. The “COVID-19 Viral Array” included 54 recombinant, purified SARS-CoV-2 proteins from commercial sources, or recombinant proteins produced in the labs of several of the authors (Peter Kim and Taia Wang, ***Supplementary Table 3***)^33^. We also included proteins or protein fragments from SARS-CoV-1, Middle East Respiratory Syndrome (MERS), nonpathogenic coronaviruses (OC43, 229E, NL63, and HKU1), Hepatitis B, Mumps, Rubella, Rubeola, Ebola, and Influenza (hemagglutinin (HA) from A/California/07/2009 H1N1).

### Array construction

Antigens were coupled to carboxylated magnetic beads (MagPlex-C, Luminex Corp.) such that each antigen was linked to beads with unique barcodes, as previously described^53, 58^. In brief, unless stated otherwise, 8 μg of each antigen or control antibody was diluted in phosphate buffered saline (PBS) and transferred to 96-well plates. Diluted antigens and control antibodies were conjugated to 1×10^6^ carboxylated magnetic beads per ID. Beads were distributed into 96-well plates (Greiner BioOne), washed and re-suspended in phosphate buffer (0.1M NaH_2_PO_4,_ pH 6.2) using a 96-well plate washer (Biotek). The bead surface was activated by adding 100 μl of phosphate buffer containing 0.5 mg 1-ethyl-3(3-dimethylamino-propyl)carbodiimide (Pierce) and 0.5 mg N-hydroxysuccinimide (Pierce). After 20 min incubation on a shaker, beads were washed and resuspended in activation buffer (0.05M 2-N-Morpholino EthaneSulfonic acid, MES, pH 5.0). Diluted antigens and control antibodies were incubated with beads for 2 hours at room temperature. Beads were washed three times in 100 μl PBS-Tween, re-suspended in 60 μ storage buffer (Blocking reagent for Enyzme Linked Immunosorbent Assay, ELISA, Roche) and stored in plates at 4°C. Immobilization of some antigens and control antibodies on the correct bead IDs was confirmed by analysis using commercially available mouse monoclonal antibodies, or antibodies specific for epitope tags such as 6X-histidine. In addition, prototype human plasma samples derived from participants with autoimmune diseases with known reactivity patterns (*e.g.* ds-DNA, Scl-70, centromere, SSA, SSB, cardiolipin, whole histones, and RNP, all purchased from ImmunoVision; also from Stanford Autoimmune Diseases Biobank, and OMRF); APS-1, IPEX, PAP, or AMI associated with anti-IFN- γ blocking antibodies; as well as normal human sera (ImmunoVision, Product # HNP-0300, certified to be nonreactive to Hep-2 cell lysates at a titer of 1:100), were used for validation.

### Array probing

Serum or plasma samples were first heat inactivated at 56°C for one hour^59^ then tested at 1:100 dilution in 0.05% PBS-Tween supplemented with 1% (w/v) bovine serum albumin (BSA) and transferred into 96-well plates in a randomized layout. The bead array was distributed into a 384-well plate (Greiner BioOne) by transfer of 5 µl bead array per well. 45 µl of the 1:100 diluted sera were transferred into the 384-well plate containing the bead array. Samples were incubated for 60 min on a shaker at room temperature. Beads were washed with 3 × 60 µl PBS-Tween on a plate washer (EL406, Biotek) and 50 µl of 1:500 diluted R-phycoerythrin (R-PE) conjugated Fcγ human IgG F(ab’)2 fragment (Jackson ImmunoResearch) was added to the 384-well plate for detection of bound human IgG. After incubation with the secondary antibody for 30 min, the plate was washed with 3 × 60 µl PBS-Tween and re-suspended in 60 µl PBS-Tween prior to analysis using a FlexMap3D^TM^ instrument (Luminex Corp.). A minimum of 100 events per bead ID were counted. Binding events were displayed as Mean Fluorescence Intensity (MFI). To ensure reproducibility and rigor, all samples were run in duplicate in each experiment. Most samples were analyzed twice using the same array run on different days, showing high concordance (mean Pearson correlation coefficient 0.94, standard deviation (SD) 0.09 for one representative dataset). Prototype autoimmune sera were also heat inactivated and compared with untreated prototype autoimmune serum on the same arrays, with similar results (data not shown).

### Anti-dsDNA ELISA

Anti-DNA ELISA was performed on calf thymus DNA (Sigma)-coated Immobilon 96 well plates that were then blocked with 1% BSA in PBS. Dilutions of patient and control plasmas in blocking buffer were incubated in the DNA-coated 96 well plates, unbound Ig was washed off, and bound IgG was measured with rabbit anti-human IgG alkaline phosphatase conjugate (Millipore). Samples were analyzed at dilutions of 1:30, 1:90, 1:270 and 1:810. Of these dilutions, 1:270 was chosen as optimal for distinguishing background binding from positive staining. Samples from five healthy donors were tested under the same conditions and an arbitrary cut-off of 2× the highest measured value at 1:270 (which was 0.390 arbitrary units) was used to distinguish positive from negative levels of binding.

### Anti-Myeloperoxidase (MPO) and Anti-Proteinase 3 (PR3) ELISAs

MPO (Cat# 708705) or purified proteinase 3 PR3 (Cat # 708700) pre-bound to the wells of microtiter plates were purchased from Inova Diagnostics (San Diego, CA). Assays were performed as recommended by the manufacturer using a dilution of 1:100 plasma in diluent (provided in the assay kit). Assay controls included strong positive, weak positive and negative samples provided by the vendor. Additional controls included samples from five healthy donors and three de-identified patients known to have clinically elevated PR3 and MPO antibody levels. Results were scored as positive or negative based upon the kit instructions.

### ANA and imaging

ANAs were performed by indirect immunofluorescence using fixed and permeabilized Hep-2 cells affixed to glass slides (Inova Diagnostics, Cat # 708100). ANAs were detected using a FITC-conjugated goat anti-human IgG antibody following vendor instructions. Samples were screened in a blinded fashion at a dilution of 1:80 with ultraviolet (UV) microscopy by clinical laboratory staff (A.G. and J.G.) who have extensive experience in the interpretation of ANA patterns. Positive and weak positive samples were tested further at 1:160 (the dilution at which ANAs are considered to be positive in the clinical lab assay, which uses the same assay kit). In addition to the kit positive and negative controls (which were included on every slide), de-identified clinical samples from patients with known clinically detectable ANAs were used.

For ANA image analysis, all images were collected with a Nikon Eclipse Ti with widefield illumination equipped with a Nikon Plan Apo VC 60× 1.4 oil objective. FITC and Evans Blue fluorescence images were collected with a Chroma dichroic/beamsplitter (part no 89402) and a Chroma quadset CoolLED300 light source, using FITC 480/30× excitation and 519/26m emission filters, as well as Cy5 640/30× excitation and 697/60m emission filters. Images were acquired with a Hamamatsu ORCA-ER B&W CCD Digital Camera controlled with Metamorph V7.10.3.390 software and 1×1 camera binning. Multiple stage positions were collected using a Ludl XY linear encoded stage and Z motor. Minimum and maximum pixel values all set to the same level on a 12-bit camera (4,096 gray levels- FITC 500 min, 2500 max, and Cy5 300 min, 1300 max), gamma set to 1, with acquisition times and light source intensities consistent for all images for comparison purposes.

### Anti-SARS-CoV-2 ELISAs

RBD ELISAs were performed as described in a previous study^60^ with several modifications. SARSCoV-2 RBD proteins (gift of Scott Hensley) were coated on the ELISA plates (Cat # 1193A15, Thomas Scientific, Swedesboro, NJ) at a final concentration of 2 μg/mL in 50 μl of 1× PBS. Serum and plasma samples were diluted 1:100 in sample dilution buffer (PBS-Tween with 1% non-fat milk powder by weight) and incubated for 1 hour at room temperature. Goat anti-human IgG-HRP (Cat# 109-035-008, Jackson ImmunoResearch Laboratories, West Grove, PA) was diluted 1:10,000 with sample dilution buffer and 50 μl of secondary antibody was added to each well. After washing 3× (PBS-Tween), plates were incubated for 0.5 hour at room temperature and washed again 3× (PBSTween). 50 μl of TMB substrate (Cat# 555214, Fisher Scientific, Waltham, MA) were applied for the color development at room temperature for 10 mins and stopped with 50 μl of 250 mM hydrochloric acid. All samples were run in duplicate. Anti-SARS-CoV-2 Nucleocapsid Assays were run with a commercially produced kit (Cat# CV3002, LifeSensors, Malvern, PA.) and performed according to the manufacturer’s instructions. Serum and plasma samples were diluted 1:100 and plates were read at OD450 nm. The RBD and nucleocapsid ELISAs were repeated to confirm the results. ELISA plates were read at OD450 nm (CLARIOStar plate reader, BMG LABTECH Inc., Cary, NC).

### Statistical Analyses

All data analysis and statistics were performed using R and various R packages^61^. For normalization, average MFI values for “bare bead” IDs were subtracted from average MFI values for antigen conjugated bead IDs. The average MFI for each antigen was calculated using samples from healthy subjects known to be uninfected with SARS-CoV-2 (all obtained before December 2019). Antibodies were considered “positive” if MFI was > 5 SD above the average MFI for HC for that antigen, and MFI was >3,000 units, a threshold which is more stringent than commonly published in related literature^21^. A less stringent 3 SD cutoff used in a Luminex assay to measure SARS-CoV-2 immunoglobulins in blood and saliva^62^ was also employed for comparison in some experiments. An example can be found in ***Supplementary Figs. 3 and 4***. ELISA and antibody number data were visualized in GraphPad Prism v.9.0.0 (86). Upon publication of this study in a peer-reviewed journal, deidentified array data will be uploaded to the Gene Expression Omnibus (GEO) database.

## Data Availability

Upon publication of this study in a peer-reviewed journal, deidentified array data will be uploaded to the Gene Expression Omnibus (GEO) database.

## Acknowledgements

**Funding**

**A.E.P.** was supported by a Stanford Maternal and Child Health Research Institute postdoctoral fellowship.

**A.F.** was funded by Stanford University and the Vice Provost for Undergraduate Education’s (VPUE) 2019-2020 Major Grant.

**A.N.** was funded by AH 06-01 (Carreras Foundation).

**C.S.** was funded by Universities Giessen and Marburg Lung Center and the German Center for Lung Research, University Hospital Gießen and Marburg (UKGM) research funding according to article 2, section 3 cooperation agreement, the Deutsche Forschungsgemeinschaft (DFG)–funded- SFB 1021 (C04), KFO 309 (P10) and SK 317/1-1 (Project number 428518790) as well as by the Foundation for Pathobiochemistry and Molecular Diagnostics.

**E.J.W.** was supported by NIH grants AI105343, AI112521, AI082630, AI201085, AI123539, and AI117950 to E.J.W. E.J.W. is also supported by the Parker Institute for Cancer Immunotherapy which supports the cancer immunology program at UPenn.

**E.L. P.** was funded by NIH UC4 DK112217 and NIH UM1-AI144288 (Autoimmunity Centers of Excellence, ACE). The ACE is a research network supported by the National Institute of Allergy and Infectious Disease (NIAID/NIH). We thank the Human Immunology Core, which assisted with COVID-19 sample processing and ELISAs, and receives infrastructure support from NIH PA30- CA016520 and NIH P30-AI0450080, and the Clinical Immunology Laboratory at the Hospital of the University of Pennsylvania.

**E.M.** was funded by Stiftung P.E. Kempkes (06/2014, 01/2016), Deutsche José Carreras Leukämie-Stiftung (18R/2016), Rhön Klinikum AG (RKA No. 64).

**H.G.** was supported by T32 HL007586.

**H.R.** was funded by the Universities Giessen Marburg Lung Center and the German Center for Lung Disease (DZL German Lung Center, no. 82DZL00502) for UGMLC.

**I.N.** was supported by UTHSC/UofM SARS-CoV-2/COVID-19 Research CORNET Award.

**K.N.** was supported by NIH Grant 5U19AI057229-17, and by philanthropic support from the Sean N Parker Center COVID-19 Research Fund.

**M.A.** was funded by the Sean N Parker Center COVID-19 Research Fund.

**M.R.** was funded by UTHSC/UofM SARS-CoV-2/COVID-19 Research CORNET Award.

**N.J.M.** was funded by NIH HL137006 and HL137915, which funded the enrollment, biosample collection, and clinical phenotyping of human subjects at Penn.

**P.J.U.** was supported by National Institute of Allergy and Infectious Diseases of the National Institutes of Health, R01 AI125197-04, NIH U01 AI150741-01S1 and the Henry Gustav Floren Trust.

**P.S.K.** was supported by the Chan Zuckerberg Biohub, and the Frank Quattrone and Denise Foderaro Family Research Fund.

**S.A.A.** was supported by 1T32AR076951-01, Allen Institute for Immunology, and the Chen Family Research Fund.

**S.C.** was supported by NIH grants UM2 AI130836, UM1 AI130839, and U19 AI104209.

**T.T.W.** was supported by Chan Zuckerberg Biohub, the Searle Scholars Program, Fast Grants, the CEND COVID Catalyst Fund, and the National Institute of Allergy and Infectious Diseases of the National Institutes of Health under award numbers R01 AI139119, U19 AI111825, and U54 CA260517.

## Author Contributions

**S.E.C., A.F., P.J.U., C.S, and E.T.L.P.** designed experiments and array panels.

**S.E.C. and S.D.** performed antigen array, ACA, and viral array production, quality control, sample runs, and data acquisition.

**W.M., J.G., A.G., A.F., K.B. and E.T.L.P.** performed and/or interpreted ANA assays.

**W.M., I.N., M.R. and E.T.L.P.** performed and/or interpreted ELISAs.

**T.W., S.C., P.A.W., A.E.P, and P.S.K.** generated key antigens, provided pre-characterized sera, and assisted with data analysis and interpretation of viral array assays.

**W.H.R.** generated key reagents for the autoantigen arrays and supervised A.H.

**M.G., S.G.** collected and processed patient blood samples.

**M.G., S.G., E.M., S.A., N.J.M., H.M.G., O.O.** collected, extracted, and analyzed clinical data.

**E.M., A.N., H.R., R.G., and S.C.** assisted in designing, recruiting, and/or following inpatient and HC cohorts.

**J.S.** provided pre-characterized COVID-19 samples Kaiser Permanente that were used to establish the array platforms and to select autoantigens for the final array.

**N.J.M., E.J.W.** designed the inpatient and HC Penn cohorts and obtained funding.

**H.M.G., O.O., N.J.M., S.A.** screened and enrolled human subjects at Penn and collected biosamples.

**M.A., N.A., and K.N.** supervised clinical data management in the CROWN clinic and performed chart reviews that enabled correlation of array results with clinical parameters.

**L.B. and M.A.** from the Stanford CROWN Research Team contributed to collection and storage of patient samples, collection of patient laboratory and clinical data, and distribution of blood samples.

**P.J. and U.S.** provided and characterized blood samples from an interferon lambda trial in patients with mild COVID-19 that were used to establish assays and compare with results from hospitalized patients.

**S.E.C., A.F., S.D., S.A., W.M., H-R.C., A.K., P.C., A.H., W.H.R., C.S., E.T.L.P. and P.J.U.** analyzed and interpreted array data.

**S.E.C., A.F., S.D., S.A., W.M., C.S., E.T.L.P. and P.J.U.** created figures and tables.

**S.E.C., A.F., R.M., C.S., E.T.L.P., and P.J.U.** wrote the paper.

**P.J.U., C.S, E.T.L.P., E.J.W., and K.N.** supervised the research.

## SUPPLEMENTARY FIGURE LEGENDS

**Supplementary Fig. 1: Anti-Nuclear Antibody (ANA) staining in University of Pennsylvania COVID-19 patient cohort. a.** IgG ANA indirect immunofluorescence results at a screening titer of 1:80, with positives confirmed at 1:160. Pie chart shows patient numbers (n=73 total), color coded by strength of staining and pattern. **b.** Analysis of ANA data in paired samples obtained on 21 patients showing the number of patients whose ANAs change over time. **d.** Images of positive ANAs from individual subjects, showing diffuse (left), nucleolar (middle) and speckled (right) staining patterns. Of note, UP01 was also weakly positive for dsDNA antibodies.

**Supplementary Fig. 2: IgG ELISAs of virus and autoantigen-binding.** Binding of IgG antibodies to SARS-CoV-2 receptor binding domain (RBD), nucleocapsid, and autoantigens (double stranded DNA (dsDNA), myeloperoxidase (MPO) and proteinase 3 (PR3)). Each symbol represents a patient (N=73). For subjects in which there were two or more time points, the D7 time point was chosen except for subject UP68, in whom the D14 time point was chosen. UP01 had a weakly positive dsDNA result. UP43 was positive for PR3.

**<u>Supplementary Figures related to Figure 1.</u>**

**Supplementary Fig. 3: Heatmap of MFI corresponding to Figure 1**.

**Supplementary Fig. 4: Validation in Kaiser Permanente cohort.** Standard deviation heatmap depicting serum IgG antibodies discovered using a first generation 26-plex bead-based protein array containing the indicated autoantigens (y-axis). Autoantigens are grouped based on disease (scleroderma, SLE/Sjögren’s, primary biliary cirrhosis (PBC)/thyroid, nucleosomes, and antigens associated with tissue inflammation. COVID-19 patients from Kaiser Permanente (left panel, n=48). HC (n=10, middle panel), and 7 prototype autoimmune disorders (right panel) are shown. Colors indicate autoantibodies whose MFI measurements are >5 SD (red), <5 or >3 SD (pink) or <3 SD (black) above the average MFI for HC. MFIs <5,000 were excluded.

**<u>Supplementary Figures related to Fig. 3:</u>**

**Supplementary Fig. 5**: **Evolution of IgG autoantibody development over time in hospitalized COVID-19 patients. a.** Heatmap using the same 53-plex bead-based protein array presented in Figure 1, containing the indicated autoantigens (x-axis). Autoantigens are grouped based on disease (scleroderma, myositis and overlap syndromes such as MCTD, SLE/Sjögren’s, gastrointestinal and endocrine disorders), DNA-associated antigens, and antigens associated with tissue inflammation or stress responses. COVID-19 patients (top panel, n=98 longitudinal COVID-19 samples, including 92 paired samples from 46 subjects and two subjects who had three available timepoints each, subject UP70 and UP71). HC (n=16, middle panel), and 8 prototype autoimmune disorders (bottom panel) are shown. **b.** Heatmap using a 41-plex array of cytokines, chemokines, growth factors, and receptors. The same samples in Panel A were also analyzed for ACA. Cytokines are grouped on the x-axis by category (interferons, interleukins, and other cytokines/growth factors/receptors). Prototype samples from patients with immunodeficiency disorders include three patients with APS1, one patient with PAP, and three patients with AMI. Colors correspond to the MFI values shown at far right.

**Supplementary Fig. 6: Standard deviation heatmap of MFI corresponding to Figure 3.**

**Supplementary Fig. 7: Number of antibodies in 21 subjects with paired D0 and D7 time point data, stratified by reactivity category. a.** Heatmap of individual subjects. Rows denote subjects and time points in days (day 0, D0 and day 7, D7). Columns denote Autoantibodies (AutoAb) and Anti-Cytokine Antibodies (ACA). Antibodies are above 5 standard deviations compared to pooled healthy controls (5 SD) or between 3 and 5 standard deviations (3 SD). White cells indicate 0 antibodies. **b.** Antibody counts at D0 and D7 stratified by antibody category. Autoantibody (AutoAb, blue) and anti-cytokine antibody (ACA, yellow) counts are shown for the same 21 subjects at Day 0, (left) and Day 7 (right). Counts were based on antibodies that were present at levels between 3 and 5 SD above the average MFI for healthy control samples. **c.** Antibodies per subject shown for different antibody level cut-offs. 5 SD = > 5 SD above the average MFI for the HC group; 3 SD = >3 and <5 SD above the average MFI for HC; In SUM, the 5 and 3 SD data were separately added for each individual. This category represents all antibodies detected at 3 SD or higher. AutoAb = autoantibodies; ACA = anti-cytokine antibodies. Horizontal red lines indicate medians. Each symbol represents a subject. P values are computed using the Wilcoxon paired rank sum test. Only D7 AutoAb vs. D7 ACA were significant (p<0.05) for the 3 SD and SUM antibody levels.

**<u>Supplementary Figures related to Figs. 4 and 5</u>:**

**Supplementary Fig. 8: Comparison between ELISA and bead-based assays. a and b.** Correlation analysis between ELISA and multiplex bead data. All samples (including multiple time points from individual subjects) were included in the correlation analysis (N=57 samples from 35 individuals). Each symbol represents a sample with the mean fluorescence intensity (MFI) vs. the optical density (OD). Spearman correlations were computed with two-tailed p-values. **c and d**. Two representative line plots for 21 patients with paired samples at D0 and D7. IgG antibodies increased over time for RBD (p=0.042, Wilcoxon rank sum test) but did not change over time for Nucleocapsid (p=0.73, NS, Wilcoxon rank sum test).

**Supplementary Fig. 9: MFI bar plots for individual autoantigens and viral antigens for the four patients with one or more newly triggered autoantibodies, corresponding to** Figure 5. Error bars represent one standard deviation of the MFI for sample replicates.

## SUPPLEMENTARY TABLES

**Supplementary Table 1.** “COVID-19 Autoantigen Array” content.

| **Autoantigen Array** |  |  |  |
| --- | --- | --- | --- |
| **Bead ID** | **Antigen** | **Vendor** | **Catalog #** |
| 1 | Bare Bead |  |  |
| 2 | Human IgG from serum | Sigma | I4506 |
| 3 | Anti-Human IgG Fc fragment Specific | Jackson | 109-005-008 |
| 4 | Anti-Human IgG (H+L) | Jackson | 109-005-003 |
| 5 | Anti-Human IgG F(ab') fragment specific | Jackson | 109-005-006 |
| 6 | Beta 2 Glycoprotein 1 | Diarect | A14901 |
| 7 | Myeloperoxidase | Diarect | A18501 |
| 8 | La/SSB | Diarect | A12801 |
| 9 | Ro52 | Diarect | A12701 |
| 10 | Proteinase 3 | Diarect | A18601 |
| 11 | Histone 1 | Immunovision | HIS-1001 |
| 12 | Histone 2A and 4 | Immunovision | HIS-1002 |
| 13 | Histone 2B | Immunovision | HIS-1003 |
| 14 | CENP B | Diarect | A12501 |
| 15 | Histone 3 | Immunovision | HIS-1004 |
| 16 | Histones | Immunovision | HIS-1000 |
| 17 | GBM | Diarect | A16801 |
| 18 | C1q | Biodesign | A90150H |
| 19 | BPI | Arotec | ATB01-02 |
| 24 | Fibrillarin | Prospec | ENZ-566 |
| 31 | U11/U12 | Origene | TP303746 |
| 38 | CENP A | Diarect | A16901 |
| 39 | EJ | Diarect | A11101 |
| 40 | HSP 70 | Stressgen | NSP-555 |
| 41 | HSP 90 | Stressgen | SPP-770 |
| 42 | Intrinsic Factor | Diarect | A16701 |
| 43 | JO1 | Diarect | A12901 |
| 44 | Ku, p70/p80 | Diarect | A17301 |
| 45 | LKM1 | Diarect | A13501 |
| 46 | MDA5 | Diarect | A30001 |
| 47 | MI-2 | Diarect | A18101 |
| 48 | PCNA | Diarect | A15401 |
| 49 | PL-12 | Diarect | A15701 |
| 50 | PL-7 | Diarect | A15601 |
| 51 | PM/ScI-75 | Diarect | A17001 |
| 52 | Nucleolin | Diarect | A19701 |
| 53 | Ribo P0 | Diarect | A14101 |
| 54 | Ribo P1 | Diarect | A14201 |
| 55 | PDC-E2 | Diarect | A17901 |
| 56 | Ribo P2 | Diarect | A14301 |
| 57 | SRP54 | Diarect | A18401 |
| 58 | PM/ScI-100 | Diarect | A16001 |
| 59 | POLR3H | Origene | TP310633 |
| 60 | PDH | Sigma | P7032 |
| 62 | Ro60 | Diarect | A17401 |
| 63 | RPP14 (Th/To) | Origene | TP760291 |
| 65 | Scl-70 | Diarect | A12401 |
| 67 | Sm/RNP | Immunovision | SRC-3000 |
| 68 | Smith | Immunovision | SMA-3000 |
| 70 | Troponin I | Prospec | PRO-1269 |
| 73 | TG | Diarect | A12201 |
| 74 | GNAL | Abnova | H00002774-P01 |
| 75 | MYH6 | Origene | TP313673 |
| 76 | TPO | Diarect | A12101 |
| 77 | ZnT8 |  |  |
| 78 | U1-snRNP 68 | Diarect | A13001 |
| 79 | U1-snRNP A | Diarect | A13101 |
| 83 | RPP25 (Th/To) | Origene | TP303538 |

**Supplementary Table 2.** “COVID-19 Cytokine Array” content.

| **Cytokine/Chemokine Array** |  |  |  |
| --- | --- | --- | --- |
| **Bead ID** | **Antigen** | **Vendor** | **Catalog #** |
| 1 | Bare Bead |  |  |
| 2 | Human IgG from serum | Sigma | I4506 |
| 3 | Anti-Human IgG Fc fragment Specific | Jackson | 109-005-008 |
| 4 | Anti-Human IgG (H+L) | Jackson | 109-005-003 |
| 5 | Anti-Human IgG F(ab') fragment specific | Jackson | 109-005-006 |
| 6 | CD74 | Prospec | PRO-1467 |
| 7 | IFN-lambda2 | Peprotech | 300-02K |
| 8 | IL-1alpha | Prospec | CYT-253 |
| 10 | ITM2B | Elabscience | PKSH032599 |
| 26 | IL-31 | Prospec | CYT-625 |
| 27 | IL-6 | Prospec | CYT-098 |
| 29 | OSM | Peprotech | 300-10 |
| 30 | IL-11 | Prospec | CYT-214 |
| 32 | IL-27 | Prospec | CYT-048 |
| 33 | CNTF | Prospec | CYT-272 |
| 34 | CT-2 | Prospec | PRO-1578 |
| 35 | LIF | Peprotech | 300-05 |
| 36 | VEGFB | Peprotech | 100-20B |
| 37 | HTRA1 | R&D | 2916-SE-020 |
| 38 | GM-CSF | Peprotech | 300-03 |
| 39 | IFN-alpha2 | R&D | 11101-2 |
| 40 | IFN-beta | Peprotech | 300-02BC |
| 41 | IFN-gamma | Peprotech | 300-02 |
| 42 | IFN-epsilon | R&D | 9667-ME-025/CF |
| 43 | IFN-lambda1 | Peprotech | 300-02L |
| 44 | IFN-lambda3 | R&D | 5259-IL-025/CF |
| 45 | IFN-omega | R&D | 11395-1 |
| 46 | IL-10 | Peprotech | 200-10 |
| 47 | IL-12p40 | Peprotech | 200-12P40 |
| 48 | IL-12p70 | Peprotech | 200-12 |
| 49 | IL-15 | Peprotech | 200-15 |
| 50 | IL-17F | Peprotech | 200-25 |
| 51 | IL-1beta | Peprotech | 200-01B |
| 52 | IL-22 | Peprotech | 200-22 |
| 55 | TNF-alpha | Peprotech | 300-01A |
| 56 | TNF-beta | Peprotech | 300-01B |
| 58 | ACE2 | Sino<br>Biological | 10108-H05H |
| 59 | Eotaxin | Peprotech | 300-21 |
| 60 | Eotaxin 2 | Peprotech | 300-33 |
| 62 | IL-17A | Peprotech | 200-17 |
| 63 | IL-33 | Peprotech | 200-33 |
| 64 | IL-7 | Peprotech | 200-07 |
| 65 | MIP-1alpha | Peprotech | 300-08 |
| 67 | PDGFBB | Peprotech | 100-14B |
| 68 | sRANK-ligand | Peprotech | 310-01C |
| 69 | TIFI-gamma | Diarect | A11001 |

**Supplementary Table 3.** “COVID-19 Viral Array” content.

| **Viral Array** |  |  |  |
| --- | --- | --- | --- |
| **Bead ID** | **Antigen** | **Vendor/Source** | **Catalog #** |
| 1 | Bare Bead |  |  |
| 2 | Human IgG from serum | Sigma | I4506 |
| 3 | Anti-Human IgG Fc fragment Specific | Jackson | 109-005-008 |
| 4 | Anti-Human IgG (H+L) | Jackson | 109-005-003 |
| 5 | Anti-Human IgG F(ab') fragment specific | Jackson | 109-005-006 |
| 9 | SARS2-FL-GCN4 | Peter Kim Lab |  |
| 16 | SARS1 RBD | Peter Kim Lab |  |
| 20 | MERS RBD | Peter Kim Lab |  |
| 24 | OC43 RBD | Peter Kim Lab |  |
| 25 | 229E-FL-GCN4 | Peter Kim Lab |  |
| 31 | NL63 RBD | Peter Kim Lab |  |
| 35 | HKU1 RBD | Peter Kim Lab |  |
| 36 | A/California/07/2009 (H1N1) HA | Peter Kim Lab |  |
| 37 | Ebola glycoprotein (Ebola GP) | Peter Kim Lab |  |
| 56 | SARS2 RBD (TW) | Taia Wang Lab |  |
| 57 | Spike S1 (TW) | Taia Wang Lab |  |
| 58 | M (TW) | Taia Wang Lab |  |
| 59 | N (TW) | Taia Wang Lab |  |
| 70 | HBSAg | MyBioSource | MBS142509 |
| 71 | Mumps | Prospec | MMP-001 |
| 72 | Rubella | Prospec | RUB-291 |
| 73 | Rubeola | MyBioSource | MBS319759 |
| 74 | NSP8 | ProSci | 97-097 |
| 75 | Spike S2 | ProSci | 10-115 |
| 76 | Orf 3a | ProSci | 10-005 |
| 77 | Orf 8 | ProSci | 10-002 |
| 78 | 3C like Proteinase | ProSci | 10-116 |
| 79 | NSP 1 | ProSci | 97-095 |
| 80 | NSP 7 | ProSci | 97-096 |
| 81 | Envelope | ProSci | 10-112 |
| 82 | ME | Sino Biological | 40598-V07E |
| 83 | NSP9 | Sino Biological | 40619-V40E |
| 92 | Plpro | Sino Biological | 40593-V07E |

**Supplementary Table 4.** Marburg University patient cohort clinical characteristics.

|  | **Marburg (N = 18)** |
| --- | --- |
| **Age [Median (IQR)]** | 66.50 (62.25-74.50) |
| **Sex [Percent; (N)]** |  |
| female | 17% (3) |
| male | 83% (15) |
| **BMI [Median (IQR)]** | 28.00 (26.17-33.35) |
| **Comorbidities [Percent; (N)]** |  |
| diabetes | 33% (6) |
| obesity | 44% (8) |
| chronic cardiac disease | 39% (7) |
| coronary heart disease | 17% (3) |
| arterial occlusive disease | 0% (0) |
| chronic obstructive pulmonary disease | 0% (0) |
| asthma | 6% (1) |
| renal insufficiency | 11% (2) |
| rheumatic disorders | 6% (1) |
| neurological neuromuscular disorders | 39% (7) |
| oncological diseases | 22% (4) |
| other immunosuppression | 22% (4) |
| **Days since start of symptoms** | ND |
| **Days since COVID19 diagnosis<sup>†</sup> [Median (IQR)]** | 9.50 (6.00-15.50) |
| **Duration of hospital stay [Median; IQR]** | 29.00; 21.50-39.75 |
| **NIH Severity score** |  |
| moderate/severe | 33% (6) |
| critical | 61% (11) |
| **Mortality** | 28% (5) |
| **Intensive Care Unit** | 78% (14) |
| **Mechanical ventilation** | 83% (15) |
| **Supplemental oxygen** | 33% (6) |
| **Therapy [Percent; (N)]** |  |
| antibiotics | 89% (16) |
| antivirals | 28% (5) |
| antimycotics | 11% (2) |
| dialysis | 33% (6) |
| Extracorporeal membrane oxygenation | 6% (1) |
| **Inflammatory Biomarkers<sup>‡</sup> [Median (IQR)]** |  |
| CRP [mg/L] | 95.20 (47.95-145.68) |
| IL-6 [pg/ml] | 95.50 (37.50-126.00) |
| *complement consumption* |  |
| C3 [g/L] | 1.40 (1.19-1.47) |
| C4 [g/L] | 0.36 (0.31-0.57) |
| CH50 [U/ml] | 48.00 (43.50-57.75) |
| AH50 [%] | 108.00 (102.20-109.00) |

<sup>†</sup>days since COVID19 diagnosis until first time point
<sup>‡</sup>first measurement during hospital stay

**Supplementary Table 5.** University of Pennsylvania patient cohort clinical characteristics.

|  | **UPenn (N = 81)** |
| --- | --- |
| **Age [Median (IQR)]** | 57.5 (34.5-80.5) |
| **Sex [Percent; (N)]** |  |
| female | 45% (36) |
| male | 55% (45) |
| **NIH Disease Severity score** |  |
| day 0 (mean, range) | 3 (2-5) |
| day 14 (mean, range) | 4 (1-7) |
| **Mortality [Percent; (N)]** | 21% (17) |
| **Thrombotic complications [Percent; (N)]** | 32% (26) |
| **Comorbidities** |  |
| Obesity | 41% (34) |
| T2DM | 40% (33) |
| Hyperlipidemia | 48% (39) |
| Chronic kidney disease | 17% (14) |

**Supplementary Table 6.** Stanford patient cohort clinical characteristics.

|  | **Stanford (N = 10)** |
| --- | --- |
| **Age [Median (IQR)]** | 44 (28-67) |
| **Sex [Percent; (N)]** |  |
| female | 60% (6) |
| male | 40% (4) |
| **Days since COVID19 diagnosis<sup>†</sup> [Median (IQR)]** | 36 (8-56) |
| **NIH Severity score** |  |
| moderate/severe | 80% (8) |
| critical | 20% (2) |
| <sup>†</sup>days since COVID19 diagnosis until first time point |  |

## References

1. Guan, W. J. et al. Clinical Characteristics of Coronavirus Disease 2019 in China. N Engl J Med 382, 1708–1720, doi:10.1056/NEJMoa2002032 (2020).

2. Chen, N. et al. Epidemiological and clinical characteristics of 99 cases of 2019 novel coronavirus pneumonia in Wuhan, China: a descriptive study. Lancet 395, 507–513, doi:10.1016/s0140-6736(20)30211-7 (2020).

3. Machhi, J. et al. The Natural History, Pathobiology, and Clinical Manifestations of SARS-CoV-2 Infections. J Neuroimmune Pharmacol 15, 359–386, doi:10.1007/s11481-020-09944-5 (2020).

4. Asfuroglu Kalkan, E. & Ates, I. A case of subacute thyroiditis associated with Covid-19 infection. J Endocrinol Invest 43, 1173–1174, doi:10.1007/s40618-020-01316-3 (2020).

5. Brancatella, A. et al. Subacute Thyroiditis After Sars-COV-2 Infection. J Clin Endocrinol Metab 105, doi:10.1210/clinem/dgaa276 (2020).

6. Carfi, A., Bernabei, R., Landi, F. & Gemelli Against, C.-P.-A. C. S. G. Persistent Symptoms in Patients After Acute COVID-19. JAMA, doi:10.1001/jama.2020.12603 (2020).

7. Patel, S. Y. et al. Anti-IFN-gamma autoantibodies in disseminated nontuberculous mycobacterial infections. J Immunol 175, 4769–4776, doi:10.4049/jimmunol.175.7.4769 (2005).

8. Woodruff, M. et al. Critically ill SARS-CoV-2 patients display lupus-like hallmarks of extrafollicular B cell activation. medRxiv, doi:10.1101/2020.04.29.20083717 (2020).

9. Woodruff, M. C., Ramonell, R. P., Lee, F. E. & Sanz, I. Broadly-targeted autoreactivity is common in severe SARS-CoV-2 Infection. medRxiv, doi:10.1101/2020.10.21.20216192 (2020).

10. Zuo, Y. et al. Prothrombotic autoantibodies in serum from patients hospitalized with COVID-19. Sci Transl Med, doi:10.1126/scitranslmed.abd3876 (2020).

11. Gruber, C. N. et al. Mapping Systemic Inflammation and Antibody Responses in Multisystem Inflammatory Syndrome in Children (MIS-C). Cell, doi:10.1016/j.cell.2020.09.034 (2020).

12. Fujii, H. et al. High levels of anti-SSA/Ro antibodies in COVID-19 patients with severe respiratory failure: a case-based review: High levels of anti-SSA/Ro antibodies in COVID-19. Clin Rheumatol 39, 3171–3175, doi:10.1007/s10067-020-05359-y (2020).

13. James, J. A. in Workshop on Post-Acute Sequelae of COVID-19.

14. Barnes, B. J. et al. Targeting potential drivers of COVID-19: Neutrophil extracellular traps. J Exp Med 217, doi:10.1084/jem.20200652 (2020).

15. Tomar, B., Anders, H. J., Desai, J. & Mulay, S. R. Neutrophils and Neutrophil Extracellular Traps Drive Necroinflammation in COVID-19. Cells 9, doi:10.3390/cells9061383 (2020).

16. Zuo, Y. et al. Neutrophil extracellular traps (NETs) as markers of disease severity in COVID- 19. medRxiv, doi:10.1101/2020.04.09.20059626 (2020).

17. Wang, E. Y. et al. Diverse Functional Autoantibodies in Patients with COVID-19. medRxiv, 2020.2012.2010.20247205, doi:10.1101/2020.12.10.20247205 (2020).

18. Price, J. V. et al. Protein microarray analysis reveals BAFF-binding autoantibodies in systemic lupus erythematosus. J Clin Invest 123, 5135–5145, doi:10.1172/JCI70231 (2013).

19. Rosenberg, J. M. et al. Neutralizing Anti-Cytokine Autoantibodies Against Interferon-α in Immunodysregulation Polyendocrinopathy Enteropathy X-Linked. Frontiers in Immunology 9, doi:10.3389/fimmu.2018.00544 (2018).

20. Ayoglu B. D. M., Crofford L, Furst D, Goldmuntz EA, Keyes-Elstein L, Mayes M, McSweeney P, Nash R, Sullivan KM, Welch B, Pinckney A, Mao R, Chung L, Khatri P, Utz PJ. Identification of unique autoantibody profiles in systemic sclerosis following hematopoietic stem cell transplantation. Sci Transl Med (2020).

21. Bastard, P. et al. Auto-antibodies against type I IFNs in patients with life-threatening COVID- 19. Science, doi:10.1126/science.abd4585 (2020).

22. Price, J. V. et al. Characterization of influenza vaccine immunogenicity using influenza antigen microarrays. PLoS One 8, e64555, doi:10.1371/journal.pone.0064555 (2013).

23. Neuman de Vegvar, H. E. et al. Microarray profiling of antibody responses against simian-human immunodeficiency virus: postchallenge convergence of reactivities independent of host histocompatibility type and vaccine regimen. J Virol 77, 11125–11138, doi:10.1128/jvi.77.20.11125-11138.2003 (2003).

24. Röltgen, K. et al. Defining the features and duration of antibody responses to SARS-CoV-2 infection associated with disease severity and outcome. Science Immunology 5, eabe0240, doi:10.1126/sciimmunol.abe0240 (2020).

25. Vlachoyiannopoulos, P. G. et al. Autoantibodies related to systemic autoimmune rheumatic diseases in severely ill patients with COVID-19. Ann Rheum Dis, doi:10.1136/annrheumdis-2020-218009 (2020).

26. Hadjadj, J. et al. Impaired type I interferon activity and inflammatory responses in severe COVID-19 patients. Science 369, 718–724, doi:10.1126/science.abc6027 (2020).

27. Galani, I.-E. et al. Untuned antiviral immunity in COVID-19 revealed by temporal type I/III interferon patterns and flu comparison. Nature Immunology 22, 32–40, doi:10.1038/s41590-020-00840-x (2021).

28. Courtney, L. P., Phelps, J. L. & Karavodin, L. M. An anti-IL-2 antibody increases serum halflife and improves anti-tumor efficacy of human recombinant interleukin-2. Immunopharmacology 28, 223–232, doi:https://doi.org/10.1016/0162-3109(94)90058-2 (1994).

29. Sato, J. et al. Enhancement of anti-tumor activity of recombinant interleukin-2 (rIL-2) by immunocomplexing with a monoclonal antibody against rIL-2. Biotherapy 6, 225–231, doi:10.1007/BF01878084 (1993).

30. Ayoglu B. D. M., Crofford L, Furst D, Goldmuntz EA, Keyes-Elstein L, Mayes M, McSweeney P, Nash R, Sullivan KM, Welch B, Pinckney A, Mao R, Chung L, Khatri P, Utz PJ. Multiplexed Autoantibody Profiles in a Systemic Sclerosis Clinical Trial Comparing Autologous Hematopoietic Stem Cell Transplantation and Cyclophosphamide [abstract]. Arthritis Rheumatol 70 (2018).

31. Rosenberg, J. M. & Utz, P. J. Protein microarrays: a new tool for the study of autoantibodies in immunodeficiency. Front Immunol 6, 138, doi:10.3389/fimmu.2015.00138 (2015).

32. Gupta, S., Nakabo, S., Chu, J., Hasni, S. & Kaplan, M. J. Association between anti-interferon-alpha autoantibodies and COVID-19 in systemic lupus erythematosus. medRxiv, 2020.2010.2029.20222000, doi:10.1101/2020.10.29.20222000 (2020).

33. Chakraborty, S. et al. Proinflammatory IgG Fc structures in patients with severe COVID-19. Nature Immunology 22, 67–73, doi:10.1038/s41590-020-00828-7 (2021).

34. Zuo, Y. et al. Neutrophil extracellular traps in COVID-19. JCI Insight 5, doi:10.1172/jci.insight.138999 (2020).

35. Nakazawa, D., Masuda, S., Tomaru, U. & Ishizu, A. Pathogenesis and therapeutic interventions for ANCA-associated vasculitis. Nat Rev Rheumatol 15, 91–101, doi:10.1038/s41584-018-0145-y (2019).

36. Narasaraju, T. et al. Neutrophilia and NETopathy as Key Pathologic Drivers of Progressive Lung Impairment in Patients With COVID-19. Frontiers in Pharmacology 11, doi:10.3389/fphar.2020.00870 (2020).

37. Magro, C. et al. Complement associated microvascular injury and thrombosis in the pathogenesis of severe COVID-19 infection: A report of five cases. Transl Res 220, 1–13, doi:10.1016/j.trsl.2020.04.007 (2020).

38. Magro, C. et al. Docket SARS CoV-2 Proteins with Cutaneous and Subcutaneous Microvasculature and their Role in the Pathogenesis of Severe COVID-10. British Journal of Dermatology (In press.).

39. Casciola-Rosen, L. et al. IgM autoantibodies recognizing ACE2 are associated with severe COVID-19. medRxiv, 2020.2010.2013.20211664, doi:10.1101/2020.10.13.20211664 (2020).

40. Takahashi, Y., Haga, S., Ishizaka, Y. & Mimori, A. Autoantibodies to angiotensin-converting enzyme 2 in patients with connective tissue diseases. Arthritis Res Ther 12, R85, doi:10.1186/ar3012 (2010).

41. Woodruff, M. C. et al. Extrafollicular B cell responses correlate with neutralizing antibodies and morbidity in COVID-19. Nat Immunol 21, 1506–1516, doi:10.1038/s41590-020-00814-z (2020).

42. Kaneko, N. et al. The Loss of Bcl-6 Expressing T Follicular Helper Cells and the Absence of Germinal Centers in COVID-19. Ssrn, 3652322, doi:10.2139/ssrn.3652322 (2020).

43. Kuri-Cervantes, L. et al. Comprehensive mapping of immune perturbations associated with severe COVID-19. Science Immunology 5, eabd7114, doi:10.1126/sciimmunol.abd7114 (2020).

44. Nielsen, S. C. A. et al. B cell clonal expansion and convergent antibody responses to SARS-CoV-2. Res Sq, doi:10.21203/rs.3.rs-27220/v1 (2020).

45. Wardemann, H. et al. Predominant autoantibody production by early human B cell precursors. Science 301, 1374–1377, doi:10.1126/science.1086907 (2003).

46. Davis, C. W. et al. Longitudinal Analysis of the Human B Cell Response to Ebola Virus Infection. Cell 177, 1566–1582.e1517, doi:10.1016/j.cell.2019.04.036 (2019).

47. Di Niro, R. et al. Salmonella Infection Drives Promiscuous B Cell Activation Followed by Extrafollicular Affinity Maturation. Immunity 43, 120–131, doi:10.1016/j.immuni.2015.06.013 (2015).

48. Jenks, S. A. et al. Distinct Effector B Cells Induced by Unregulated Toll-like Receptor 7 Contribute to Pathogenic Responses in Systemic Lupus Erythematosus. Immunity 49, 725–739.e726, doi:10.1016/j.immuni.2018.08.015 (2018).

49. Lau, C. M. et al. RNA-associated autoantigens activate B cells by combined B cell antigen receptor/Toll-like receptor 7 engagement. Journal of Experimental Medicine 202, 1171–1177, doi:10.1084/jem.20050630 (2005).

50. Marshak-Rothstein, A. & Rifkin, I. R. Immunologically active autoantigens: the role of toll-like receptors in the development of chronic inflammatory disease. Annu Rev Immunol 25, 419–441, doi:10.1146/annurev.immunol.22.012703.104514 (2007).

51. Fujinami, R. S., von Herrath, M. G., Christen, U. & Whitton, J. L. Molecular mimicry, bystander activation, or viral persistence: infections and autoimmune disease. Clin Microbiol Rev 19, 80–94, doi:10.1128/cmr.19.1.80-94.2006 (2006).

52. Fujinami, R. S. & Oldstone, M. B. Amino acid homology between the encephalitogenic site of myelin basic protein and virus: mechanism for autoimmunity. Science 230, 1043–1045, doi:10.1126/science.2414848 (1985).

53. Degn, S. E. et al. Clonal Evolution of Autoreactive Germinal Centers. Cell 170, 913–926.e919, doi:10.1016/j.cell.2017.07.026 (2017).

54. Sng, J. et al. AIRE expression controls the peripheral selection of autoreactive B cells. Sci Immunol 4, doi:10.1126/sciimmunol.aav6778 (2019).

55. Ayoglu, B. et al. Anoctamin 2 identified as an autoimmune target in multiple sclerosis. Proc Natl Acad Sci U S A 113, 2188–2193, doi:10.1073/pnas.1518553113 (2016).

56. Walter, J. E. et al. Broad-spectrum antibodies against self-antigens and cytokines in RAG deficiency. J Clin Invest 126, 4389, doi:10.1172/JCI91162 (2016).

57. Furman, D. et al. Apoptosis and other immune biomarkers predict influenza vaccine responsiveness. Mol Syst Biol 9, 659, doi:10.1038/msb.2013.15 (2013).

58. Ayoglu, B. et al. Anoctamin 2 identified as an autoimmune target in multiple sclerosis. Proceedings of the National Academy of Sciences 113, 2188–2193 (2016).

59. Amanat, F. et al. A serological assay to detect SARS-CoV-2 seroconversion in humans. Nature Medicine 26, 1033–1036, doi:10.1038/s41591-020-0913-5 (2020).

60. Kuri-Cervantes, L. et al. Immunologic perturbations in severe COVID-19/SARS-CoV-2 infection. bioRxiv, doi:10.1101/2020.05.18.101717 (2020).

61. Team, R. C. R: A language and environment for statistical computing. R Foundation for Statistical Computing, Vienna, Austria. (2013).

62. Randad, P. R. et al. COVID-19 serology at population scale: SARS-CoV-2-specific antibody responses in saliva. medRxiv (2020).

