## Supplementary Figures 1-9 for "New-Onset IgG Autoantibodies in Hospitalized Patients with COVID-19"

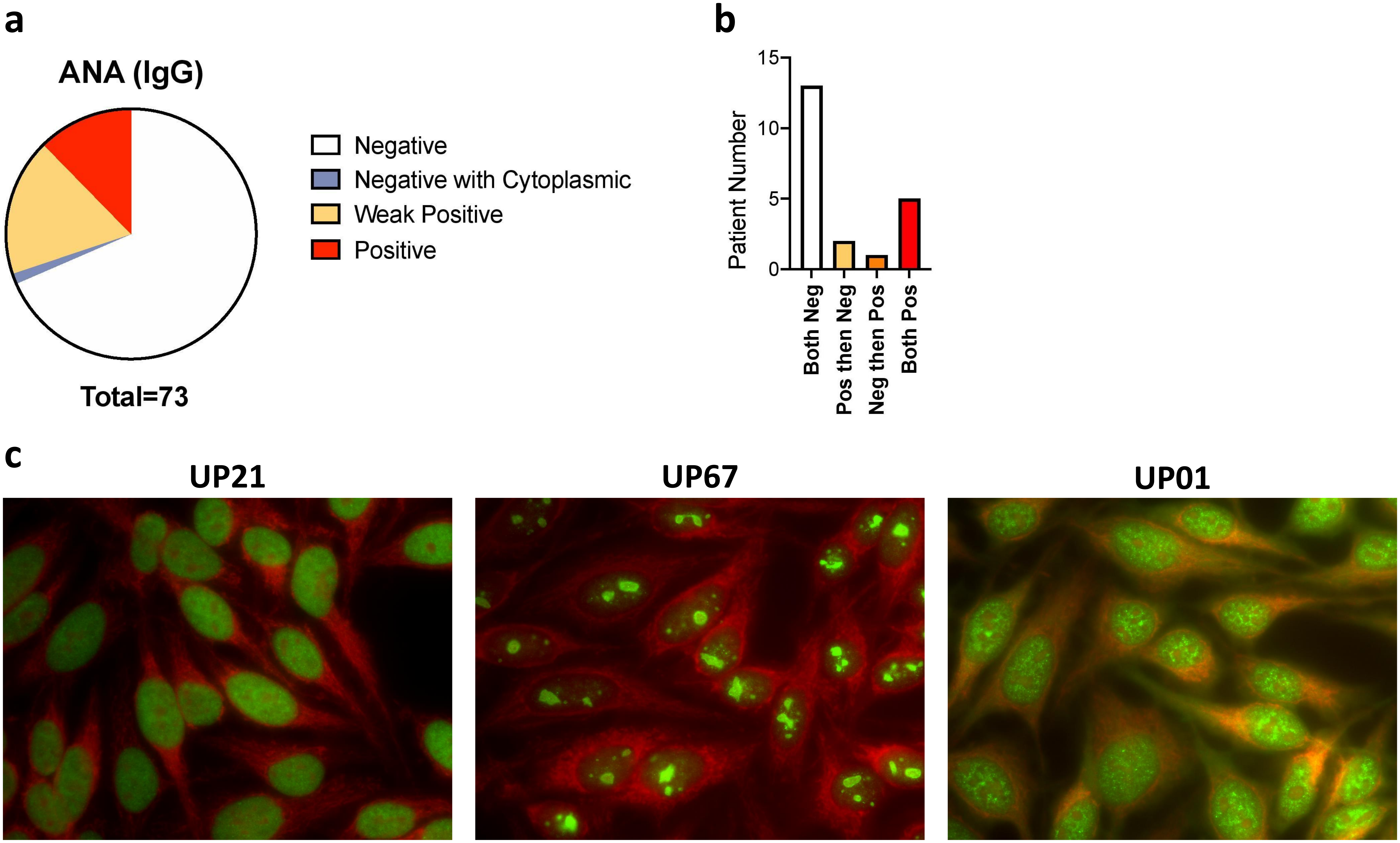


**Supplementary Fig. 1: Anti-Nuclear Antibody (ANA) staining in University of Pennsylvania COVID-19 patient cohort.** **a.** IgG ANA indirect immunofluorescence results at a screening titer of 1:80, with positives confirmed at 1:160. Pie chart shows patient numbers (n=73 total), color coded by strength of staining and pattern. **b.** Analysis of ANA data in paired samples obtained on 21 patients showing the number of patients whose ANAs change over time. **d.** Images of positive ANAs from individual subjects, showing diffuse (left), nucleolar (middle) and speckled (right) staining patterns. Of note, UP01 was also weakly positive for dsDNA antibodies.

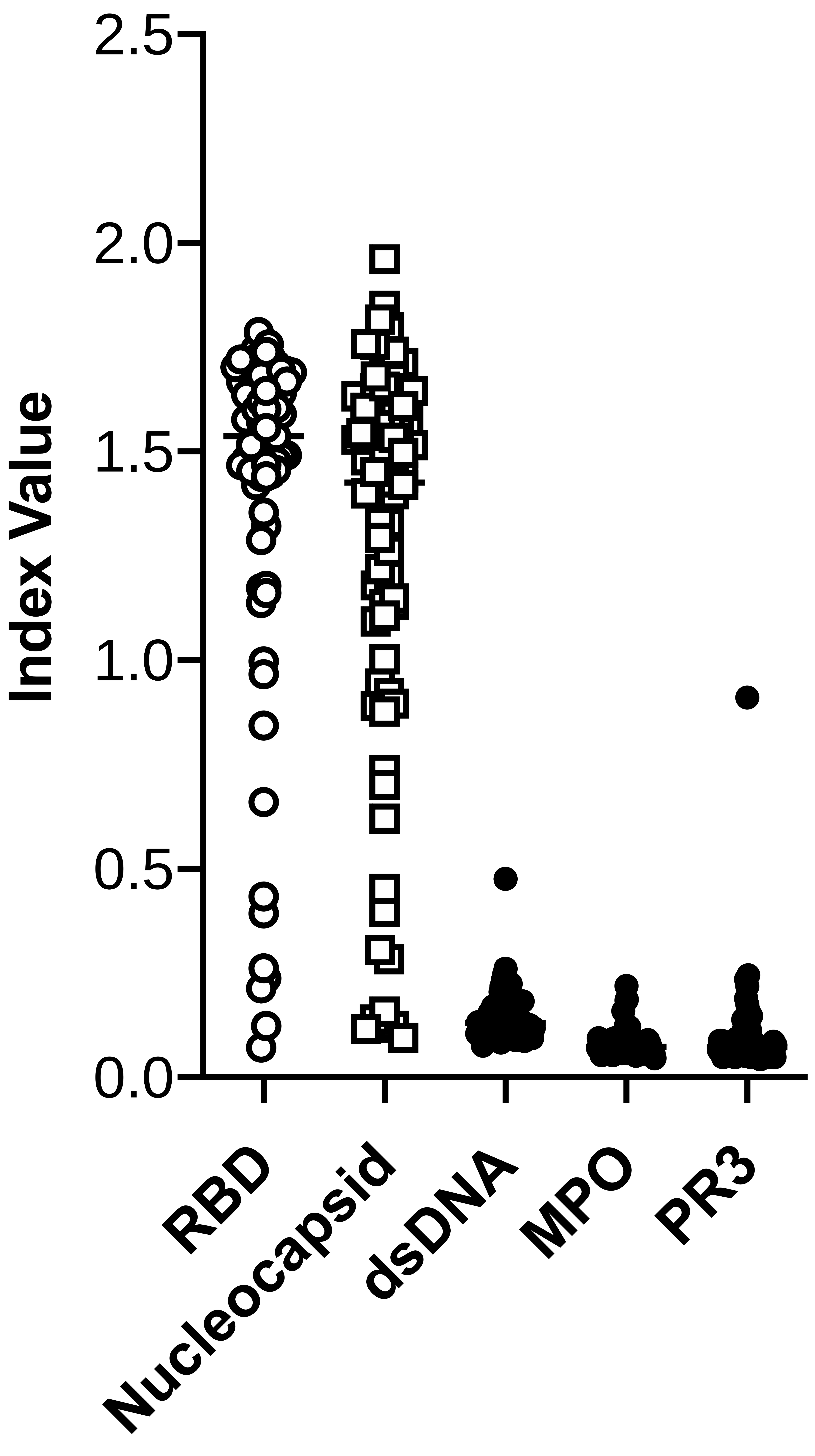


**Supplementary Fig. 2: IgG ELISAs of virus and autoantigen-binding.** Binding of IgG antibodies to SARS-CoV-2 receptor binding domain (RBD), nucleocapsid, and autoantigens (double stranded DNA (dsDNA), myeloperoxidase (MPO) and proteinase 3 (PR3)). Each symbol represents a patient (N=73). For subjects in which there were two or more time points, the D7 time point was chosen except for subject UP68, in whom the D14 time point was chosen. UP01 had a weakly positive dsDNA result. UP43 was positive for PR3.

### Supplementary Fig. 3: Heatmap of MFI corresponding to Figure 1.

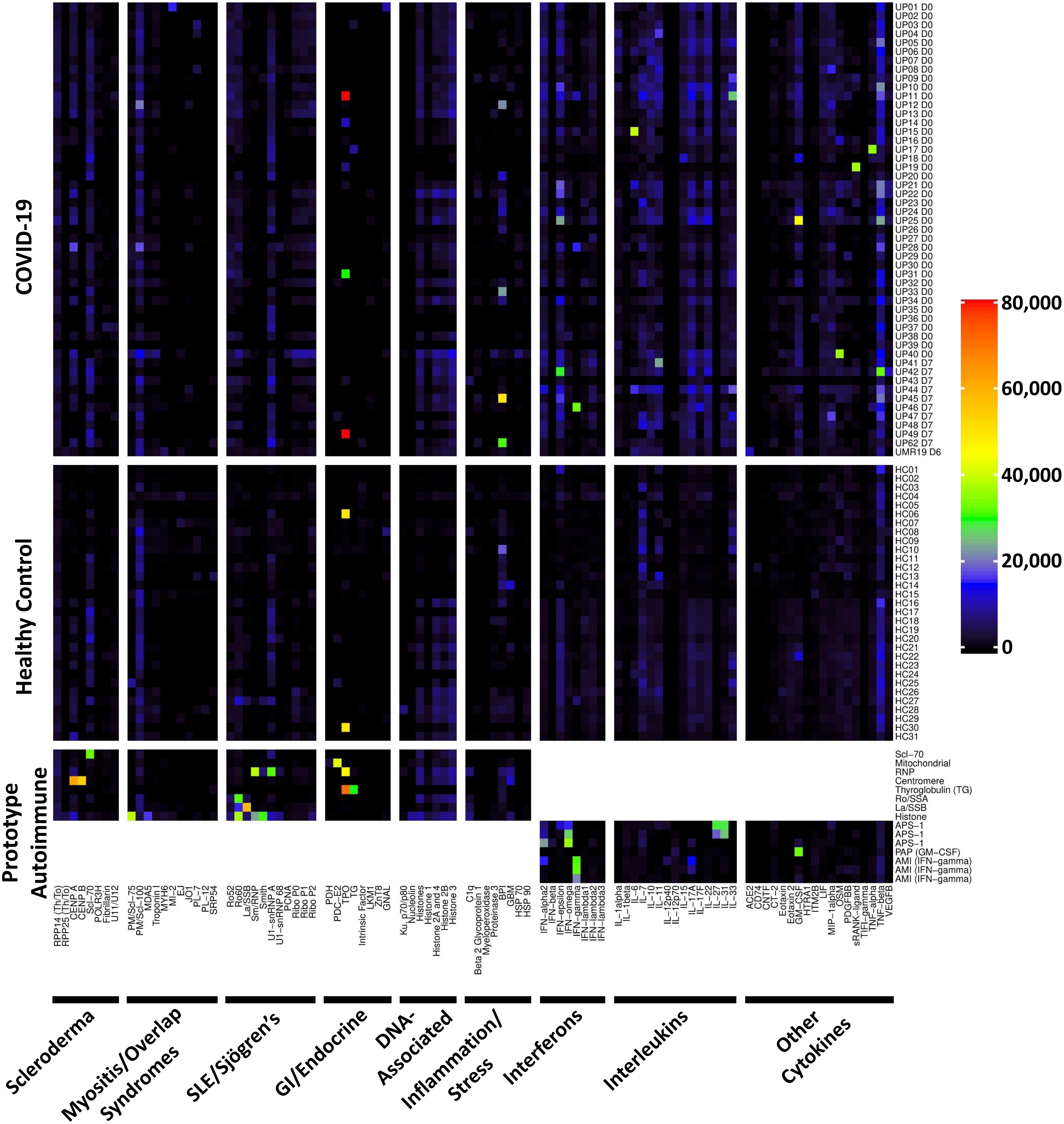

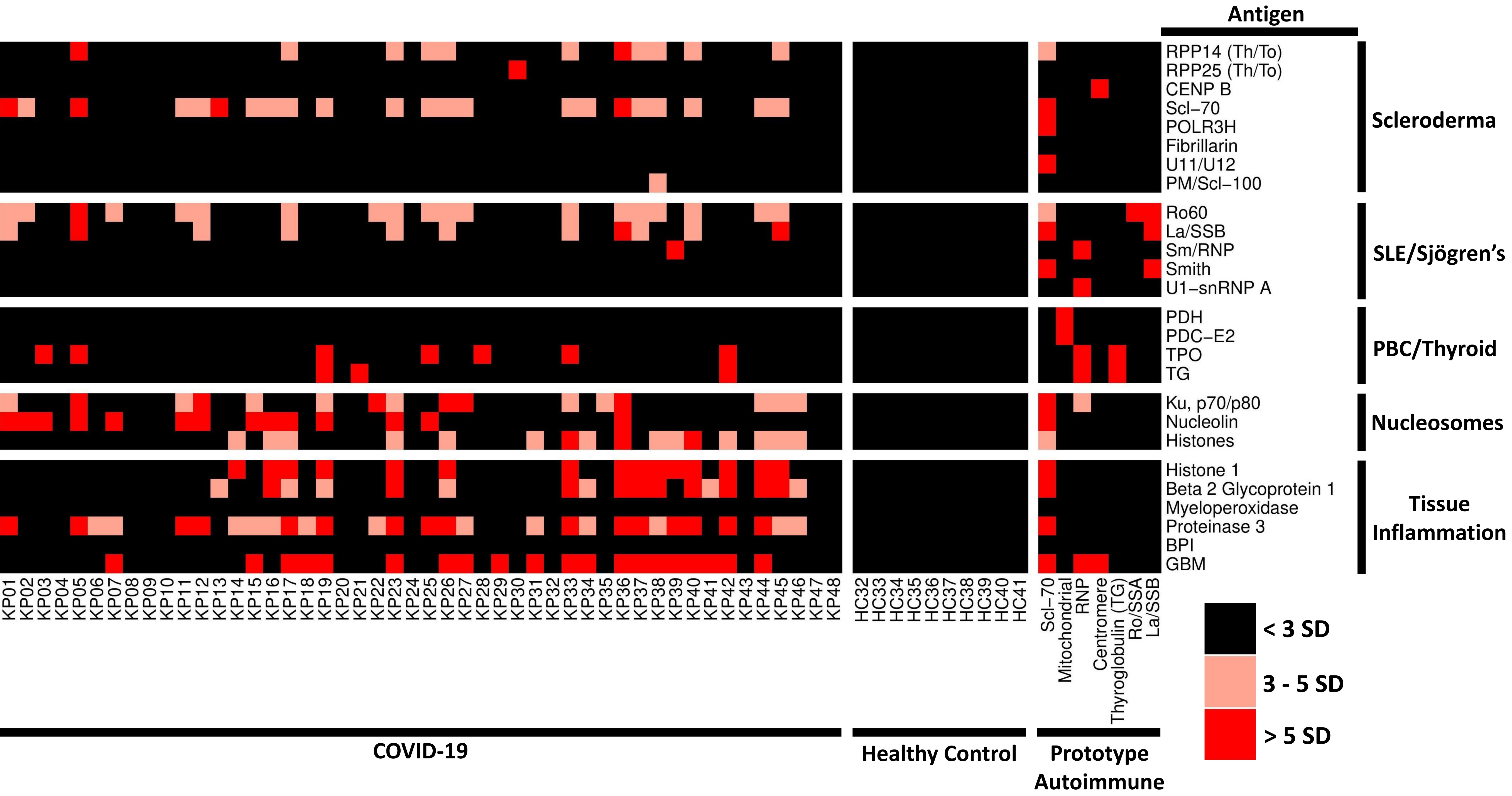


**Supplementary Fig. 4: Validation in Kaiser Permanente cohort.** Standard deviation heatmap depicting serum IgG antibodies discovered using a first generation 26-plex bead-based protein array containing the indicated autoantigens (y-axis). Autoantigens are grouped based on disease (scleroderma, SLE/Sjögren's, primary biliary cirrhosis (PBC)/thyroid, nucleosomes, and antigens associated with tissue inflammation. COVID-19 patients from Kaiser Permanente (left panel, n=48). HC (n=10, middle panel), and 7 prototype autoimmune disorders (right panel) are shown. Colors indicate autoantibodies whose MFI measurements are >5 SD (red), <5 or >3 SD (pink) or <3 SD (black) above the average MFI for HC. MFIs <5,000 were excluded.

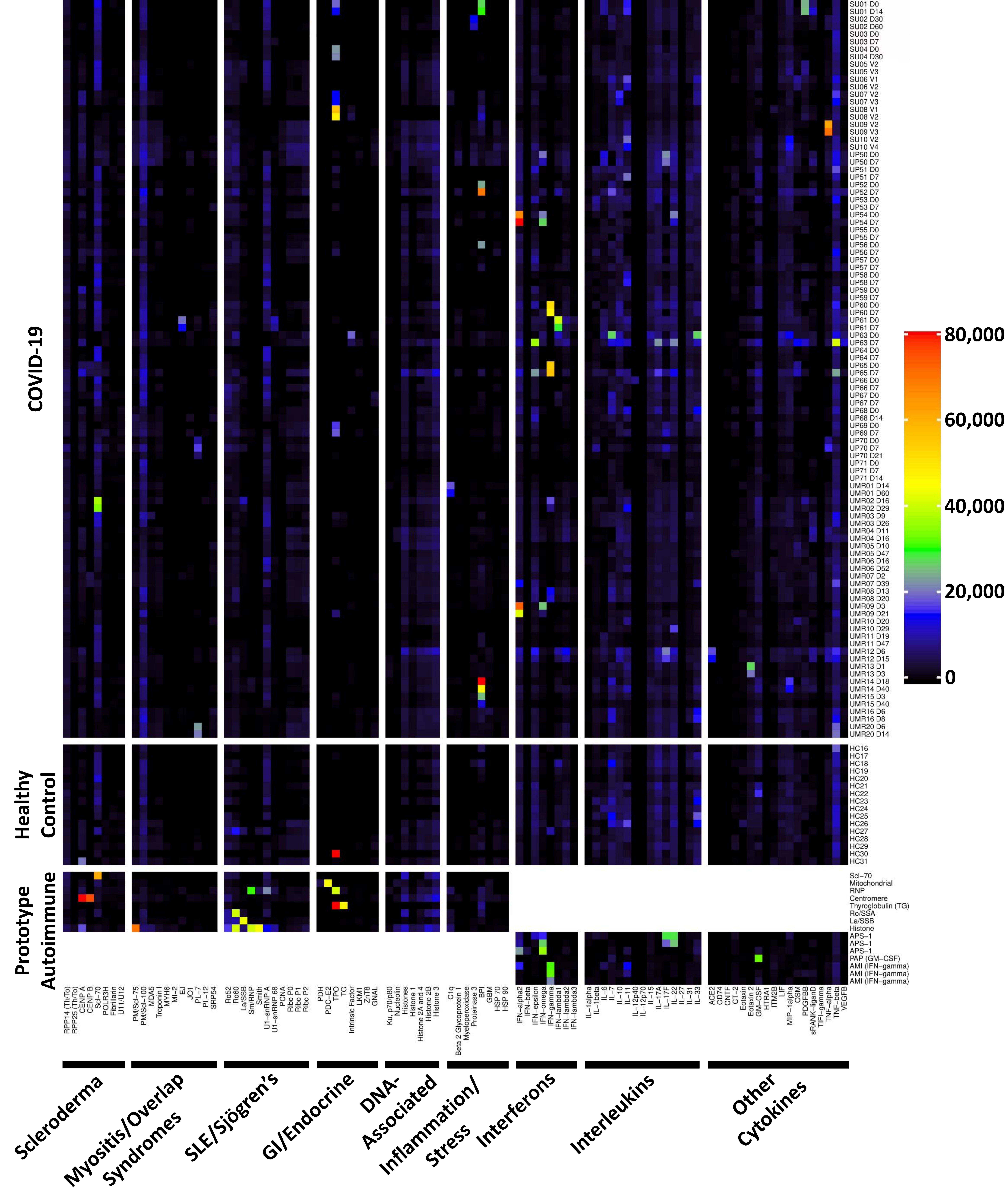

COVID-19
Healthy Control
Prototype Autoimmune
SU01 D0
SU01 D14
SU02 D30
SU02 D60
SU03 D0
SU03 D7
SU04 D0
SU04 D30
SU05 V2
SU05 V3
SU06 V1
SU06 V2
SU07 V2
SU07 V3
SU08 V1
SU08 V2
SU09 V2
SU09 V3
SU10 V2
SU10 V4
UP50 D0
UP50 D7
UP51 D0
UP51 D7
UP52 D0
UP52 D7
UP53 D0
UP53 D7
UP54 D0
UP54 D7
UP55 D0
UP55 D7
UP56 D0
UP56 D7
UP57 D0
UP57 D7
UP58 D0
UP58 D7
UP59 D0
UP59 D7
UP60 D0
UP60 D7
UP61 D0
UP61 D7
UP63 D0
UP63 D7
UP64 D0
UP64 D7
UP65 D0
UP65 D7
UP66 D0
UP66 D7
UP67 D0
UP67 D7
UP68 D0
UP68 D14
UP69 D0
UP69 D7
UP70 D0
UP70 D7
UP70 D21
UP71 D0
UP71 D7
UP71 D14
UMR01 D14
UMR01 D60
UMR02 D16
UMR02 D29
UMR03 D9
UMR03 D26
UMR04 D11
UMR04 D16
UMR05 D10
UMR05 D47
UMR06 D16
UMR06 D52
UMR07 D2
UMR07 D39
UMR08 D13
UMR08 D20
UMR09 D3
UMR09 D21
UMR10 D20
UMR10 D29
UMR11 D19
UMR11 D47
UMR12 D6
UMR12 D15
UMR13 D1
UMR13 D3
UMR14 D18
UMR14 D40
UMR15 D3
UMR15 D40
UMR16 D6
UMR16 D8
UMR20 D6
UMR20 D14
HC16
HC17
HC18
HC19
HC20
HC21
HC22
HC23
HC24
HC25
HC26
HC27
HC28
HC29
HC30
HC31
Scl−70
Mitochondrial
RNP
Centromere
Thyroglobulin (TG)
Ro/SSA
La/SSB
Histone
APS−1
APS−1
APS−1
PAP (GM−CSF)
AMI (IFN−gamma)
AMI (IFN−gamma)
AMI (IFN−gamma)
80,000
60,000
40,000
20,000
0
RPP14 (Th/To)
RPP25 (Th/To)
CENP A
CENP B
Scl−70
POLR3H
Fibrillarin
U11/U12
PM/Scl−75
PM/Scl−100
MDA5
Troponin I
MYH6
MI−2
EJ
JO1
PL−7
PL−12
SRP54
Ro52
Ro60
La/SSB
Sm/RNP
Smith
U1−snRNP A
U1−snRNP 68
PCNA
Ribo P0
Ribo P1
Ribo P2
PDH
PDC−E2
TPO
TG
Intrinsic Factor
LKM1
ZnT8
GNAL
Ku, p70/p80
Nucleolin
Histones
Histone 1
Histone 2A and 4
Histone 2B
Histone 3
C1q
Beta 2 Glycoprotein 1
Myeloperoxidase
Proteinase 3
BPI
GBM
HSP 70
HSP 90
IFN−alpha2
IFN−beta
IFN−epsilon
IFN−omega
IFN−gamma
IFN−lambda1
IFN−lambda2
IFN−lambda3
IL−1alpha
IL−1beta
IL−6
IL−7
IL−10
IL−11
IL−12p40
IL−12p70
IL−15
IL−17A
IL−17F
IL−22
IL−27
IL−31
IL−33
ACE2
CD74
CNTF
CT−2
Eotaxin
Eotaxin 2
GM−CSF
HTRA1
ITM2B
LIF
MIP−1alpha
OSM
PDGFBB
sRANK−ligand
TIFI−gamma
TNF−alpha
TNF−beta
VEGFB
Scleroderma
Myositis/Overlap Syndromes
SLE/Sjögren's
GI/Endocrine
DNA-Associated
Inflammation/Stress
Interferons
Interleukins
Other Cytokines

**Supplementary Fig. 5**: **Evolution of IgG autoantibody development over time in hospitalized COVID-19 patients. a.** Heatmap using the same 53-plex bead-based protein array presented in Figure 1, containing the indicated autoantigens (x-axis). Autoantigens are grouped based on disease (scleroderma, myositis and overlap syndromes such as MCTD, SLE/Sjögren's, gastrointestinal and endocrine disorders), DNA-associated antigens, and antigens associated with tissue inflammation or stress responses. COVID-19 patients (top panel, n=98 longitudinal COVID-19 samples, including 92 paired samples from 46 subjects and two subjects who had three available timepoints each, subject UP70 and UP71). HC (n=16, middle panel), and 8 prototype autoimmune disorders (bottom panel) are shown. **b.** Heatmap using a 41-plex array of cytokines, chemokines, growth factors, and receptors. The same samples in Panel A were also analyzed for ACA. Cytokines are grouped on the x-axis by category (interferons, interleukins, and other cytokines/growth factors/receptors). Prototype samples from patients with immunodeficiency disorders include three patients with APS1, one patient with PAP, and three patients with AMI. Colors correspond to the MFI values shown at far right.

**Supplementary Fig. 6: Standard deviation heatmap of MFI corresponding to Figure 3.**

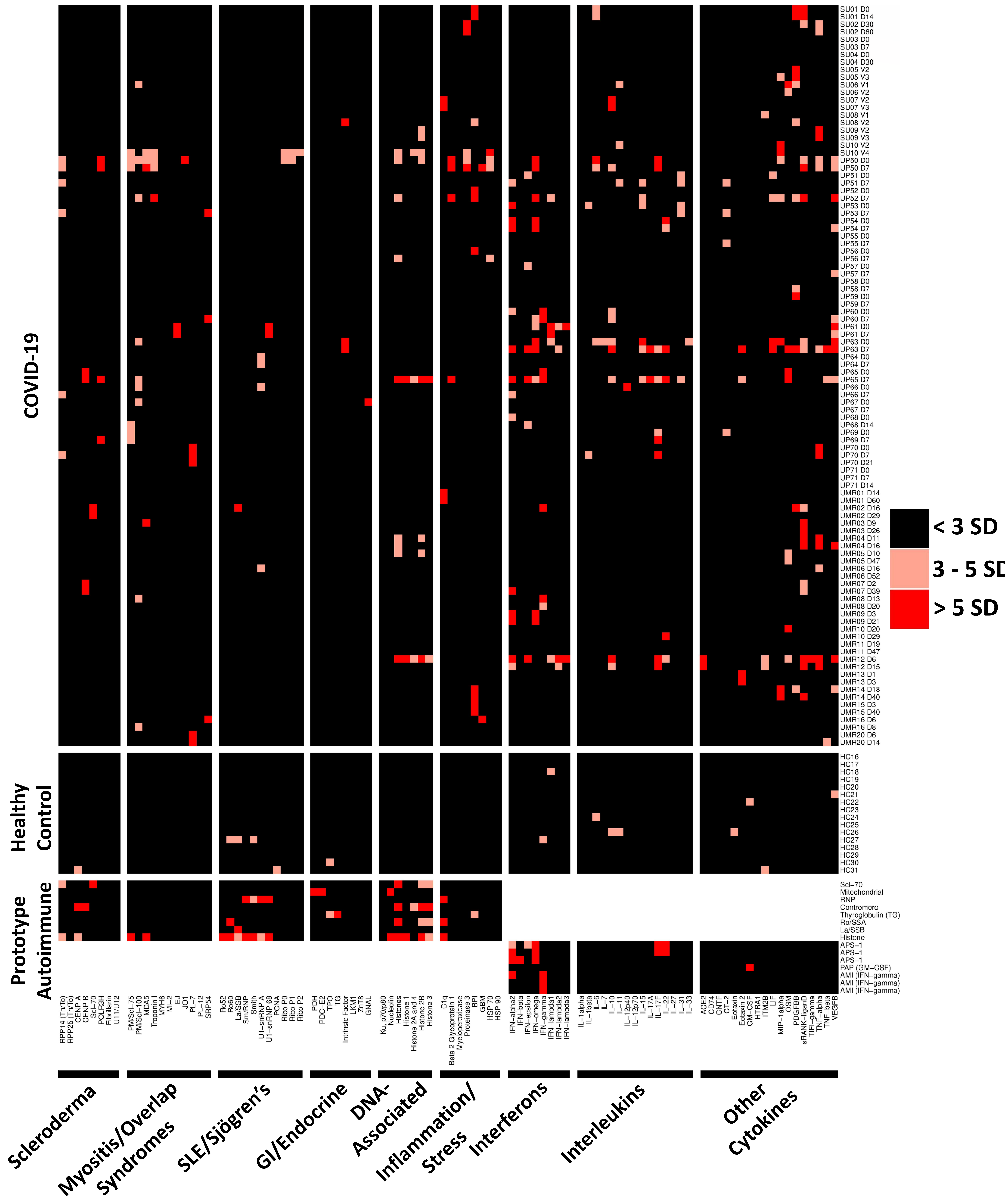

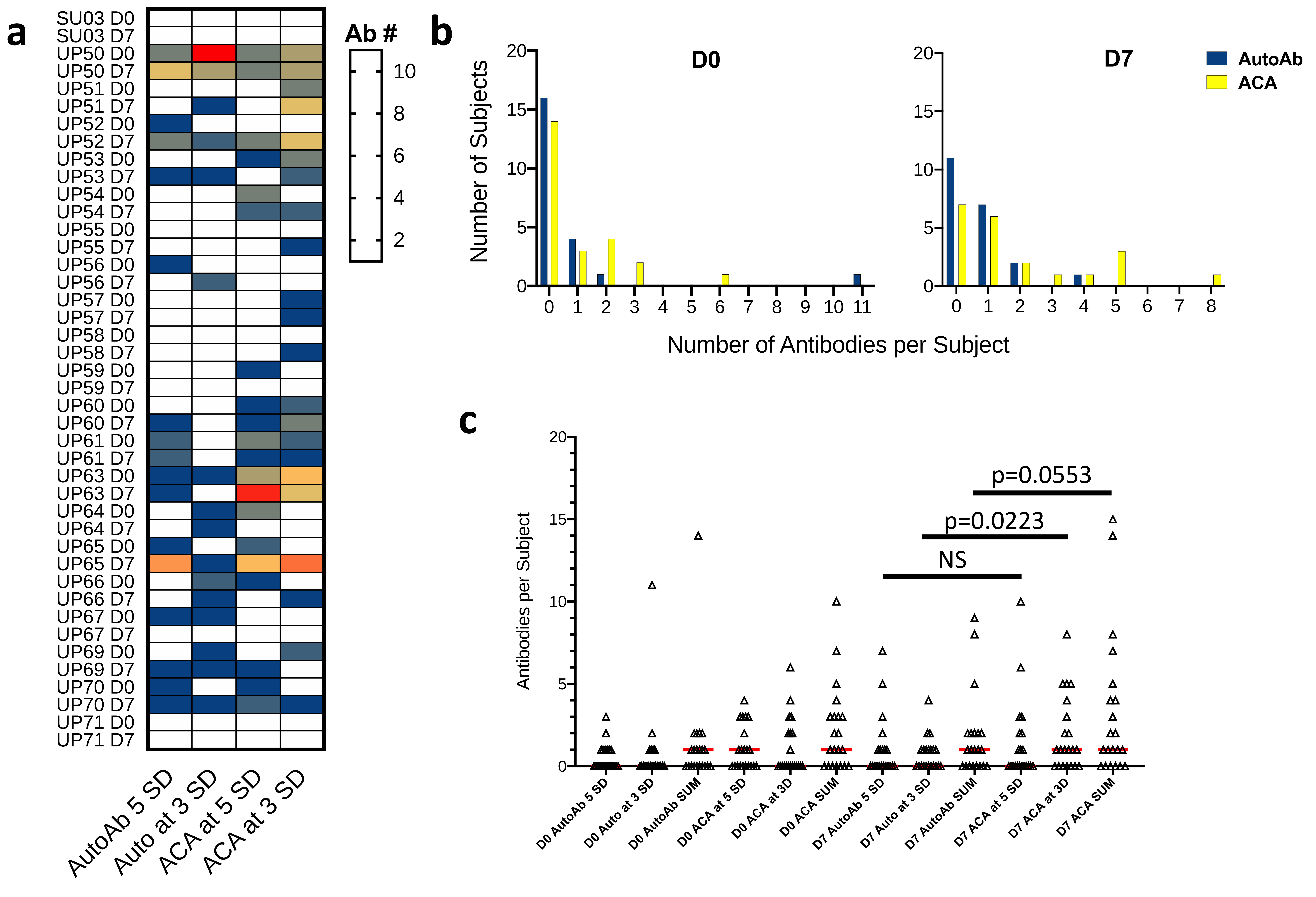

a
SU03 D0
SU03 D7
UP50 D0
UP50 D7
UP51 D0
UP51 D7
UP52 D0
UP52 D7
UP53 D0
UP53 D7
UP54 D0
UP54 D7
UP55 D0
UP55 D7
UP56 D0
UP56 D7
UP57 D0
UP57 D7
UP58 D0
UP58 D7
UP59 D0
UP59 D7
UP60 D0
UP60 D7
UP61 D0
UP61 D7
UP63 D0
UP63 D7
UP64 D0
UP64 D7
UP65 D0
UP65 D7
UP66 D0
UP66 D7
UP67 D0
UP67 D7
UP69 D0
UP69 D7
UP70 D0
UP70 D7
UP71 D0
UP71 D7
AutoAb 5 SD
Auto at 3 SD
ACA at 5 SD
ACA at 3 SD
Ab #
10
8
6
4
2
b
D0
D7
AutoAb
ACA
Number of Subjects
Number of Antibodies per Subject
c
Antibodies per Subject
p=0.0553
p=0.0223
NS
D0 AutoAb 5 SD
D0 Auto at 3 SD
D0 AutoAb SUM
D0 ACA at 5 SD
D0 ACA at 3D
D0 ACA SUM
D7 AutoAb 5 SD
D7 Auto at 3 SD
D7 AutoAb SUM
D7 ACA at 5 SD
D7 ACA at 3D
D7 ACA SUM

**Supplementary Fig. 7: Number of antibodies in 21 subjects with paired D0 and D7 time point data, stratified by reactivity category. a.** Heatmap of individual subjects. Rows denote subjects and time points in days (day 0, D0 and day 7, D7). Columns denote Autoantibodies (AutoAb) and Anti-Cytokine Antibodies (ACA). Antibodies are above 5 standard deviations compared to pooled healthy controls (5 SD) or between 3 and 5 standard deviations (3 SD). White cells indicate 0 antibodies. **b.** Antibody counts at D0 and D7 stratified by antibody category. Autoantibody (AutoAb, blue) and anti-cytokine antibody (ACA, yellow) counts are shown for the same 21 subjects at Day 0, (left) and Day 7 (right). Counts were based on antibodies that were present at levels between 3 and 5 SD above the average MFI for healthy control samples. **c.** Antibodies per subject shown for different antibody level cut-offs. 5 SD = > 5 SD above the average MFI for the HC group; 3 SD = >3 and <5 SD above the average MFI for HC; In SUM, the 5 and 3 SD data were separately added for each individual. This category represents all antibodies detected at 3 SD or higher. AutoAb = autoantibodies; ACA = anti-cytokine antibodies. Horizontal red lines indicate medians. Each symbol represents a subject. P values are computed using the Wilcoxon paired rank sum test. Only D7 AutoAb vs. D7 ACA were significant ($p<0.05$) for the 3 SD and SUM antibody levels.

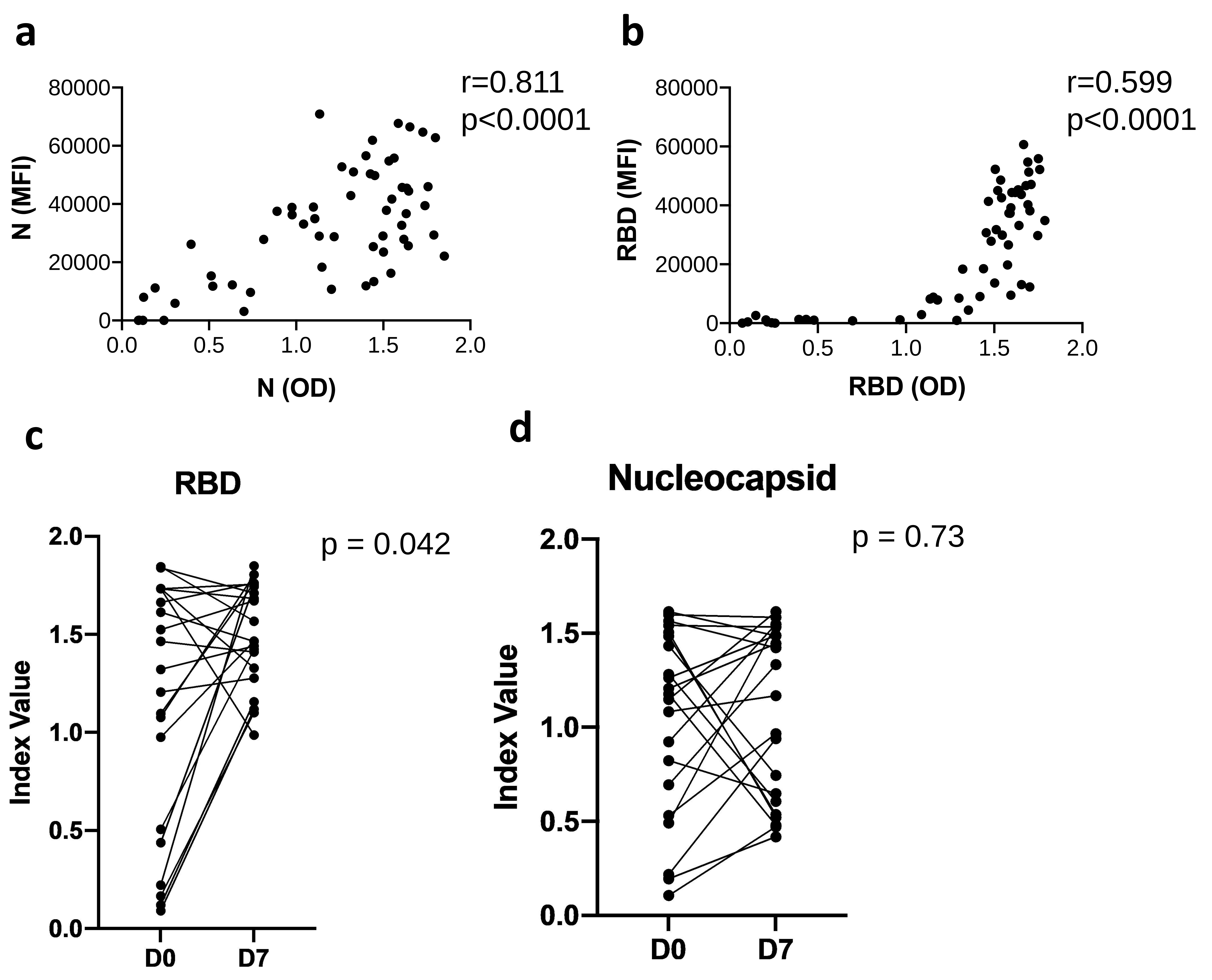

a
r=0.811
p<0.0001
N (MFI)
N (OD)
b
r=0.599
p<0.0001
RBD (MFI)
RBD (OD)
c
RBD
p = 0.042
Index Value
D0
D7
d
Nucleocapsid
p = 0.73
Index Value
D0
D7

**Supplementary Fig. 8: Comparison between ELISA and bead-based assays. a and b.** Correlation analysis between ELISA and multiplex bead data. All samples (including multiple time points from individual subjects) were included in the correlation analysis (N=57 samples from 35 individuals). Each symbol represents a sample with the mean fluorescence intensity (MFI) vs. the optical density (OD). Spearman correlations were computed with two-tailed p-values. **c and d**. Two representative line plots for 21 patients with paired samples at D0 and D7. IgG antibodies increased over time for RBD ($p=0.042$, Wilcoxon rank sum test) but did not change over time for Nucleocapsid ($p=0.73$, NS, Wilcoxon rank sum test).

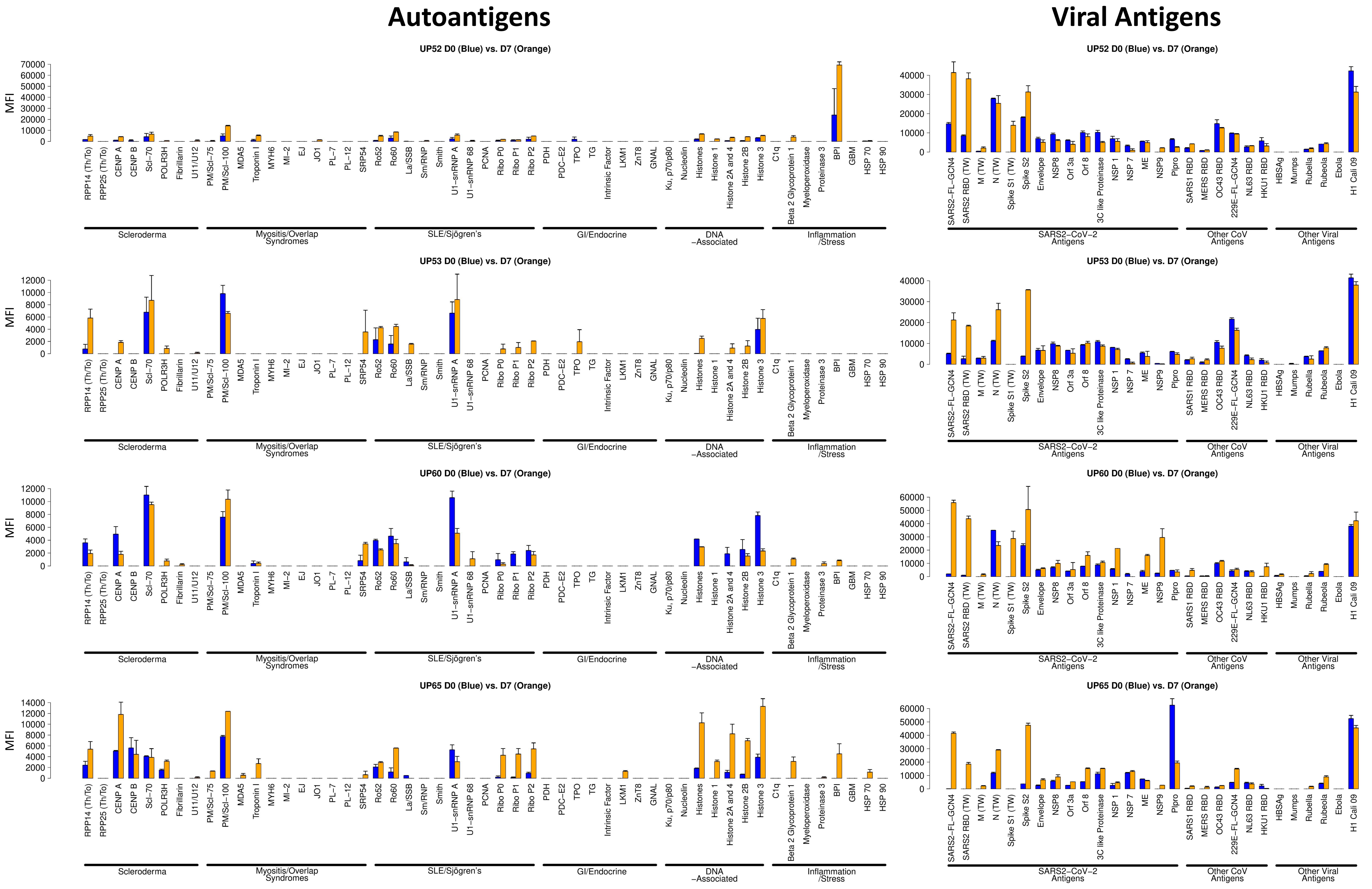


**Supplementary Fig. 9: MFI bar plots for individual autoantigens and viral antigens for the four patients with one or more newly triggered autoantibodies, corresponding to Figure 5.** Error bars represent one standard deviation of the MFI for sample replicates.
